# Interpretable Multimodal Machine Learning Model for Predicting Health Risks of Patients with Heart Failure

**DOI:** 10.64898/2026.01.12.26343905

**Authors:** Rachel Chae, Jiandong Zhou, Oscar Hou In Chou, Bingqing Yang, Haolin Pu, Gary Tse, Bernard Man Yung Cheung, Tingting Zhu, Josip Car, Lei Lu

## Abstract

Heart failure (HF) is one of the major causes of morbidity and mortality globally, necessitating accurate tools for health outcome prediction and risk stratification. In this study, we propose an interpretable multimodal machine learning framework integrating four clinical data modalities (i.e., demographics, medications, laboratory tests, and electrocardiograms [ECGs]) to predict 30-day all-cause mortality and hospital readmission in HF patients. Using clinical data from 2,868 HF patients across 43 local hospitals in Hong Kong, we trained and evaluated ten machine learning models for HF risk prediction, with the best performing model achieving an area under the receiver operating characteristic curve (AUC) of 0.881 for mortality and 0.709 for readmission. Notably, laboratory tests and ECG features dominate predictive power, and their combination alone yielded near-optimal results (AUC: 0.872), suggesting that these two modalities may be adequate for effective risk prediction in resource-constrained settings. The SHapley Additive exPlanations (SHAP) analysis identified serum albumin, high-sensitivity troponin I, lactate dehydrogenase, and QT interval dispersion as key predictors. Feature redundancy analysis further revealed strong correlations within laboratory tests and ECG features, suggesting opportunities for model simplification. To the best of our knowledge, this is the first study that comprehensively evaluates diverse configurations of four data modalities for HF risk prediction through ablation analysis, quantifying the marginal gains of each data modality and their combinations. Our findings demonstrate that interpretable multimodal machine learning model can enhance risk prediction in HF patients, supporting personalized management and scalable deployment across diverse healthcare settings.

## 1. Introduction

Cardiovascular diseases (CVDs) are the leading cause of mortality and morbidity globally, affecting over 500 million people and accounting for approximately 20.5 million deaths in 2021 [1], [2]. Among CVDs, heart failure (HF) is a particularly challenging clinical condition due to its progressive nature and high rates of adverse outcomes. Notably, up to one in four heart failure patients is readmitted to the hospital within 30 days of discharge [3], reflecting both the clinical complexity of the condition and its substantial strain on healthcare systems. In the United States alone, unplanned readmissions within 30 days of discharge contribute to more than $17 billion annual healthcare care expenditures [4]. These challenges underscore the critical need for accurate risk stratification tools to enable early identification of high-risk HF patients and facilitate timely, targeted interventions.

HF is a complex clinical syndrome characterized by the inability of the heart to pump sufficient blood to meet the body’s metabolic demands, often resulting from underlying conditions such as coronary artery disease, hypertension, or cardiomyopathy [5]. It is typically diagnosed and stratified through a combination of clinical history, physical examination, biomarker testing (e.g., B-type natriuretic peptide, BNP) and imaging modalities such as echocardiography to assess left ventricular ejection fraction and structural abnormalities [6], [7]. For example, the MAGGIC (Meta-Analysis Global Group in Chronic Heart Failure) is a simple yet powerful clinical method of risk stratification for HF patients with preserved ejection fraction [8]. The MAGGIC was developed to predict 1- and 3-year mortality in HF patients using 13 clinical variables, including age, sex, body mass index, systolic blood pressure, ejection fraction, creatinine, current smoker, diabetes mellitus, chronic obstructive pulmonary disease, New York Heart Association class, HF duration >18 months, *β*-blocker use, and angiotensin-converting enzyme inhibitor use [8], [9]. Previous studies showed that the MAGGIC risk model achieved promising performance in predicting 1-year mortality at population level [10], and it was useful for optimizing selection of patients with HF who need specialist care [11]. However, these traditional approaches rely on static and manually curated variables that may not capture the full complexity or dynamic progression of HF risks. Furthermore, predictive scores such as MAGGIC were derived from historical cohorts and often do not incorporate real-time patient conditions that could enhance early risk detection and personalized treatment.

Recent advances in machine learning (ML) have enabled more sophisticated modeling of complex and heterogeneous data associated with HF risks. Various ML methods have been explored for the prediction of HF risk, such as logistic regression [12], random forests [13], deep neural networks [14], transformer-based models [15], and large language models [16]. Among these, gradient boosting models (GBMs), particularly XGBoost (eXtreme Gradient Boosting), have gained significant attention due to their strong predictive performance and interpretability when combined with SHAP (SHapley Additive exPlanations) [17]–[19]. Leveraging these ML frameworks, various data sources have been utilized to support HF diagnosis, risk stratification, and disease management. For example, echocardiography is commonly used for screening and guiding treatment decisions [20], [21]. Electronic health record (EHR) and administrative data have enabled the identification of hospitalized HF cases [12], [18] and prediction of 30-day outcomes [22], [23]. Electrocardiograms (ECGs) have facilitated HF detection via deep learning [14], and identification of digital biomarkers related to atrial fibrillation and mortality [24]. Cardiac magnetic resonance (CMR) imaging has further demonstrated utility in phenotyping and predicting long-term outcomes, particularly in patients with suspected cardiac sarcoidosis [25].

While these studies demonstrate the potential of various data sources for HF risk prediction [14], [15], [20], [21], [24], they are largely limited to single-modality inputs, which capture only a partial view of the patient’s condition. For example, EHR data may lack real-time physiological dynamics [15]; ECG provides rich electrophysiological signals but it can be easily affected by measurement noises and artifacts [26]; while imaging modalities are often costly and not routinely available at scale [25]. As a result, single-modality models risk missing critical interactions across biological systems and may underperform in real-world settings where data completeness varies [27]. These limitations underscore the need for integrative, multimodal approaches that combine complementary information from diverse sources, such as demographics, medications, laboratory tests, and ECG features, to achieve more robust, generalizable, and clinically actionable risk predictions.

The integration of multimodal data has emerged as a promising strategy for HF risk prediction in recent studies. For example, multimodality imaging has been used to refine the prognosis in patients with HF and preserved ejection fraction [28]; combinations of echocardiographic and laboratory data have been applied to predict risk in ischemic cardiomyopathy and HF with preserved ejection fraction [29]; 12-lead multimodal ECG signals have been used for the early diagnosis of HF [30]; and integration of clinical notes with structured tabular data has been shown to improve prognostic accuracy for hospital outcomes in patients with HF [31]. These efforts reflect a growing recognition that combining diverse data sources can capture complementary aspects of disease severity and progression. Despite these advances, many existing studies on multimodal HF risk prediction face critical limitations: (*i*) they often rely on small, single-center cohorts with limited statistical power and generalizability; (*ii*) when ECG or other physiological signals are included, they are typically treated as supplementary features without rigorous analysis to quantify their incremental value over robust EHR-only baselines; (*iii*) there is a lack of interpretability-focused investigation into cross-modal interactions, such as how specific abnormal features modify the risk conveyed by laboratory biomarkers, which could provide mechanistic insights and guide clinical action. As a result, clear evidence is still lacking regarding when and for whom multimodal models outperform the strong EHR-only baselines, and which modality interactions drive those improvements in predictive performance.

This study aims to address this gap by developing and evaluating a multimodal ML framework for predicting 30-day outcomes in HF patients using a comprehensive dataset. Then, we identify the most predictive clinical and physiological markers through SHAP-based explainability, and assess the incremental value of integrating diverse data modalities. Furthermore, we explore feature interactions and redundancy to inform future model simplification and clinical deployment. To the best of our knowledge, this is the first study to perform a systematic ablation analysis across four modalities, i.e., demographics, medications, laboratory tests, and ECG features, to quantify the relative contribution of each to HF risk prediction. This comprehensive evaluation not only highlights the importance of multimodal integration but also provides practical guidance for feature selection and model design in real-world settings.

## 2. Methods

### 2.1. Study design

An overview of our study design is illustrated in Figure 1. First, a dataset comprising four types of medical records and ECG data was used to derive diverse features, which were paired with records of 30-day mortality and 30-day hospital readmission of HF patients. The labeled features were then stratified into training, validation, and test sets. The training and validation sets were used to train several baseline models, and their performance on the test set was compared. Finally, the best-performing model was selected for further analysis of its predictive performance, model explainability, and selection of important features.

**Figure 1.**
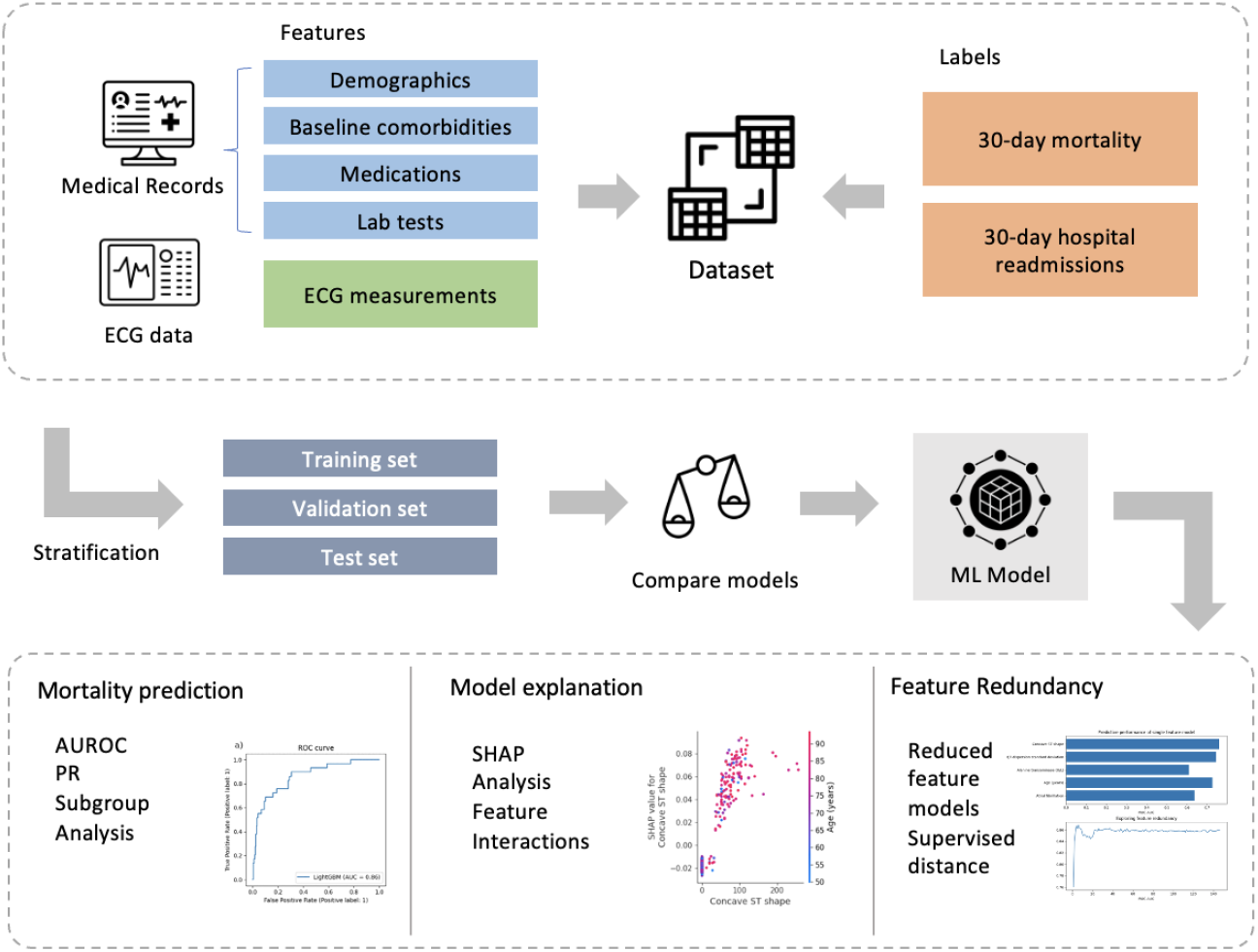
Flowmap of model training, validation, and analysis for HF risk prediction.

### 2.2. Data sets and cohorts

This study uses four types of structured electronic health records and tabulated ECG data derived from the Clinical Data Analysis and Reporting System (CDARS), an integrated territory-wide database managed by the Hong Kong Hospital Authority that consolidates patient information from 43 public hospitals and associated outpatient clinics across Hong Kong. Although data extraction, cleaning, and analysis were performed by investigators affiliated with a single tertiary centre in Hong Kong, the underlying cohort reflects a multicentre public-hospital population captured in CDARS rather than a single-centre cohort. HF patients were identified from CDARS based on eligibility criteria and the availability of at least one 12-lead baseline ECG recorded, and a total of 2,868 HF patients were included in the dataset, with a median age of 77.37 years and an interquartile range (IQR) of 66.89 to 84.28 years (Table 1).

**Table 1.**
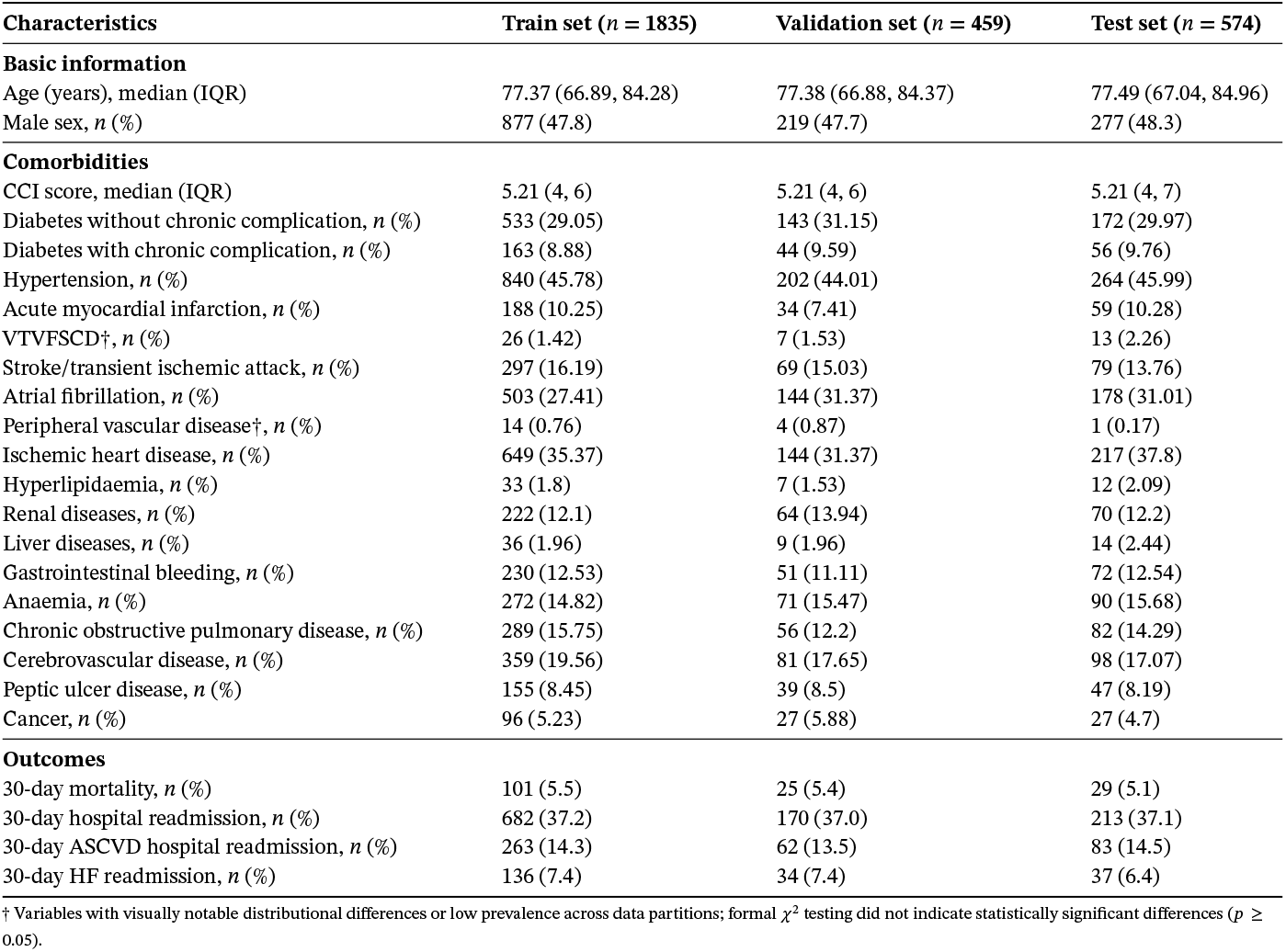
Distribution of patient characteristics and health outcomes in the training, validation, and test sets.

The dataset included coded medical records encompassing patient demographics, baseline comorbidities, healthcare utilization history, medication prescriptions, and laboratory tests (e.g., complete blood counts, renal/liver function tests, lipid and glucose tests). To avoid information leakage, all predictors were defined using pre-baseline observation windows only, where the ECG measurement date was designated as the baseline (time zero) for cohort entry and feature extraction. Comorbidities were defined from diagnosis codes recorded within the 5 years prior to the index admission. Medications were defined from prescription records within the 12 months prior to the index ECG. Laboratory values were extracted from measurements obtained prior to the index ECG, using the closest available pre-baseline test within a 6-month window when multiple results were available. Healthcare utilization variables (e.g., prior hospitalizations and emergency visits) were captured only if they occurred prior to the index admission. Outcomes were defined as 30-day all-cause mortality and 30-day all-cause hospital readmission following discharge from the index admission. No variables recorded after the index ECG time or after discharge from the index admission were used as model inputs.

The dataset also contained ECG measurements from patients upon admission. The raw ECG data were processed and tabulated into a total of 78 features, such as heart rate variability (HRV), P-wave durations, and QT intervals. The details of ECG signal processing and feature extraction can be found in our previous research [32]. Combined with features from medical records, a total of 2868 samples with 146 features were used as tabulated inputs to machine learning models. Details of clinical characteristics of HF patients and the list of extracted features can be found in ***Supplementary*** file.

Information on patient health outcomes after hospital discharge was provided in terms of mortality and hospital readmission within 30 days after initial admission. Mortality data was sourced from the Hong Kong Death Registry, an official government registry containing death records for all Hong Kong citizens, which is linked to the CDARS. Although additional information on the cause of mortality and hospital readmission was also provided (e.g., 30-day CVD mortality, 30-day atrial fibrillation readmission), the models were trained using all-cause 30-day mortality and 30-day hospital readmission as the prediction targets, as these are widely used indicators in evaluating health outcomes of HF patients [33], [34].

The dataset arose from the doctoral studies of the author Dr. Gary Tse (G.T.) at the Graduate School, The Chinese University of Hong Kong. G.T. obtained ethics approval from the Chinese University of Hong Kong-New Territories East Cluster Clinical Research Ethics Committee for studies related to the use of ECGs for risk stratification (Approval numbers: 2019.338, 2019.361, and 2019.422), which were used towards his doctoral thesis. The requirement for informed consent was waived because of the retrospective design and a lack of patient contact. Furthermore, G.T. received written confirmation from the M.D. Subcommittee of the Graduate School of the Chinese University of Hong Kong for the use of the data. The study was conducted in accordance with the Declaration of Helsinki. This cohort was obtained as subset of a previously conducted retrospective registry study of adult patients hospitalized for heart failure between 2005 and 2019 from a single tertiary centre [35]. The registry has been used to conduct single centre [32], [36], [37], multi-centre studies [38], and territory-wide cross-cluster studies from the city [39].

### 2.3. Train-test dataset split

Of the entire dataset, 20% of the data (574 samples) were hold out for testing. The remaining data were divided into training (64% of total dataset, 1835 samples) and validation sets (16% of total dataset, 459 samples). The train-test split was stratified by demographic characteristics (e.g., sex, age), as well as two health outcomes (e.g., 30-day mortality and 30-day hospital readmission rates). Due to the imbalance between positive and negative samples for one of the labels of interest (30-day mortality), stratification was performed when splitting the dataset into training, validation, and testing sets to ensure that the positive samples were well distributed in the subsets used for training and testing. Statistical tests were performed to ensure that there was an even distribution of health outcomes from the overall dataset. A chi-square test was performed to ensure that the sex and health outcomes of training, testing, and validation sets did not differ significantly from each other, and a two-sample independent t-test was performed to check that the age distributions of the datasets did not differ significantly from each other.

### 2.4. Model development and evaluation

Several ML model architectures and parameters were explored in the process of identifying the best performing model for predicting health outcomes of HF patients.

We acknowledge that previous studies have shown promising performance of tree-based models over deep learning models in analyzing tabular data, particularly with respect to model tuning, interpretability, and prediction on moderate or small size datasets [40]. Therefore, we selected a range of baseline models for developing the risk prediction model and for feature importance interpretation. These models include XGBoost, LightGBM, Random Forest, Logistic Regression, and neural networks. The models were developed on the training set and optimized with hyperparameter tuning. Baseline models that do not natively support missing values (e.g., logistic regression, support vector machines, and neural networks) required explicit imputation during training. Missing values for these baseline models were handled using multivariate imputation by chained equations (MICE), as implemented within the AutoGluon preprocessing pipeline. In contrast, the final LightGBM model relied on its intrinsic handling of missing values during tree construction, and no explicit imputation or post-baseline data processing was performed prior to training.

The set of model parameters that resulted in the highest AUC score (LightGBM) was chosen as the best performing model. The resulting models for each of the model architectures were evaluated based on their performance on the test sets. AUC was used as the primary discrimination metric. Given class imbalance (particularly for 30-day mortality), we additionally report precision–recall curves and the area under the precision–recall curve (AUPRC), alongside threshold-dependent metrics (F1, precision, recall) computed using operating thresholds selected on the validation set and applied to the test set.

### 2.5. Model explanation

Feature importance was analyzed to generate model explanation and visualize variables that were the most important in shaping model predictions. We used the SHAP technique to determine the importance of clinical variables in structured tabular data [41]. Higher SHAP values indicate greater importance of specific features in influencing the model’s prediction performance, while lower SHAP values suggest a less significant influence. Using SHAP techniques provides valuable insight into the relationships between input features and prediction outcomes, allowing a better understanding of the model’s decision-making process.

Feature interactions were explored in several ways throughout this study. First, the feature importance ranking from SHAP analysis was used to identify the features that are the most impactful on the model’s predictive performance. The top features’ interactions with model outcomes, as well as their interactions with each other, were explored by mapping the SHAP values of individual features. Then, the model was retrained on subsets of features, either individual features identified as impactful in the SHAP analysis or categories of features (e.g., demographics, laboratory tests, ECG measurements), and the predictive performance of each model was also compared. This comprehensive analysis provided insight into which modalities contributed clinically significant information and were the most instrumental component in model prediction.

## 3. Results

### 3.1. Patient characteristics

In the dataset, 155 out of 2868 samples (5.4%) were reported for mortality within 30 days of discharge. The train-test split was stratified on demographic characteristics (e.g., sex and age) as well as health outcomes (e.g., 30-day mortality and 30-day hospital readmission) to ensure even distribution of labels across training and testing data. The patient characteristics of the training, validation, and test sets are summarized in Table 1. Statistical tests (chi-square test and two-sample independent t-test) were performed to ensure that the distribution of demographic characteristics and health outcomes of train, validation, and test sets did not differ significantly from each other.

### 3.2. Baseline model performances

A total of ten baseline models were evaluated for 30-day mortality prediction, including Logistic Regression, Random Forest, Decision Tree, XGBoost, LightGBM, Support Vector Machine (SVM), three neural-network variants (NeuralNetwork, NNAttention, NNResidual), and TabNet. AutoGluon TabularPredictor [42] was used as the primary framework for baseline screening and standardized preprocessing where applicable, while several models were trained using standalone implementations outside AutoGluon.

Logistic Regression, Random Forest, XGBoost, and LightGBM models were trained within AutoGluon, which provides a unified pipeline for preprocessing, missing-value handling, and model optimization using scikit-learn and gradient boosting backends. Decision Tree, SVM, neural-network variants, and TabNet were trained separately using their respective reference implementations, while maintaining identical training, validation, and test splits to ensure comparability across models.

Neural network baselines included a feed-forward multilayer perceptron (NeuralNetwork) with two hidden layers and ReLU activations, trained using the Adam optimizer with dropout regularization to mitigate overfitting. Two architectural variants were additionally evaluated to assess the effect of network design choices: NNResidual incorporates residual (skip) connections to improve optimization stability and gradient flow in deeper representations, while NNAttention augments the feed-forward architecture with a self-attention mechanism to enable flexible modeling of feature interactions.

TabNetClassifier [44], a deep learning architecture employing sequential attention masks for tabular feature selection, was also evaluated using default configurations, providing a representative comparison against attention-based tabular models.

All models were evaluated on the same held-out test set. Threshold-dependent metrics (F1 score, precision, and recall) were computed using operating thresholds selected on the validation set and applied unchanged to the test set. Table 2 summarizes model discrimination (AUC) and operating-point performance. Among the evaluated approaches, gradient-boosted tree models demonstrated the strongest discrimination performance. LightGBM achieved the highest AUC, followed closely by XGBoost, and was therefore selected as the primary model for subsequent analysis.

**Table 2.** Baseline model performance for predicting 30-day mortality.

| Model | AUC | F1 | Precision | Recall |
| --- | --- | --- | --- | --- |
| LogisticRegression | 0.7363 | 0.2185 | 0.1444 | 0.4483 |
| RandomForest | 0.8480 | 0.2647 | 0.2308 | 0.3103 |
| DecisionTree | 0.6939 | 0.3467 | 0.2826 | 0.4483 |
| XGBoost | 0.8508 | 0.3214 | 0.2169 | 0.6207 |
| SVMClassifier | 0.7610 | 0.2319 | 0.2000 | 0.2759 |
| NeuralNetwork | 0.6276 | 0.1628 | 0.1228 | 0.2414 |
| NNAttention | 0.7011 | 0.2041 | 0.1449 | 0.3448 |
| NNResidual | 0.7075 | 0.2697 | 0.2000 | 0.4138 |
| TabNetClassifier | 0.6904 | 0.1983 | 0.1304 | 0.4138 |
| **LightGBM** | **0.8566** | **0.3248** | **0.2159** | **0.6552** |

Note: The F1, precision, and recall are computed on the test set using operating thresholds selected on the validation set.

Having identified LightGBM as the best overall performer, we ran a cross-validated hyperparameter sweep on the training set using AUC as the selection metric, then evaluated on the held-out test set. Figure 2 shows validation and test ROC curves for the two final models: 30-day mortality (validation AUC = 0.93; test AUC = 0.88) and 30-day readmission (validation AUC = 0.73; test AUC = 0.71). Operating thresholds were chosen on the validation ROC using Youden’s J (maximize TPR-FPR) [43] and then fixed for test evaluation. Applying these same thresholds to the test set produced the confusion matrices in Figure 3. For prediction of 30-day mortality (thr = 0.10), we observed TN (True Negative) = 381, FP (False Positive) = 164, FN (False Negative) = 3, TP (True Positive) = 26, reflecting a sensitivity-favored operating point suitable for ruling out risk. For prediction of 30-day readmission (thr = 0.41), we observed TN = 202, FP = 159, FN = 44, TP = 169, consistent with the lower discriminative signal in this endpoint. As shown in Figure 4, we further presented the precision–recall (PR) curves and the Area Under the PR Curve (AUPRC) for the LightGBM model in predicting 30-day mortality and 30-day hospital readmission. Solid lines show test-set performance (mortality AUPRC = 0.33; readmission AUPRC = 0.57), and dashed lines show validation-set performance (mortality AUPRC = 0.39; readmission AUPRC = 0.61).

**Figure 2.**
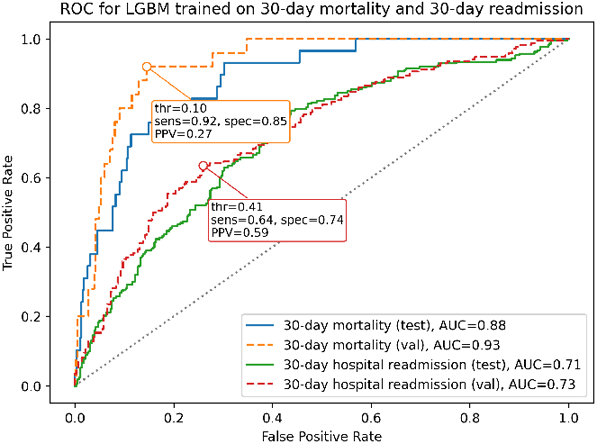
ROC curves for the LightGBM model on predicting 30-day mortality and 30-day hospital readmission. Solid lines show test performance (mortality AUC = 0.88; readmission AUC = 0.71); dashed lines show validation performance (mortality AUC = 0.93; readmission AUC = 0.73). Callouts mark validation-selected operating thresholds (thr) via Youden’s J approach [43] (mortality thr = 0.10 and readmission thr = 0.41). The dotted diagonal indicates chance performance (AUC = 0.5).

**Figure 3.**
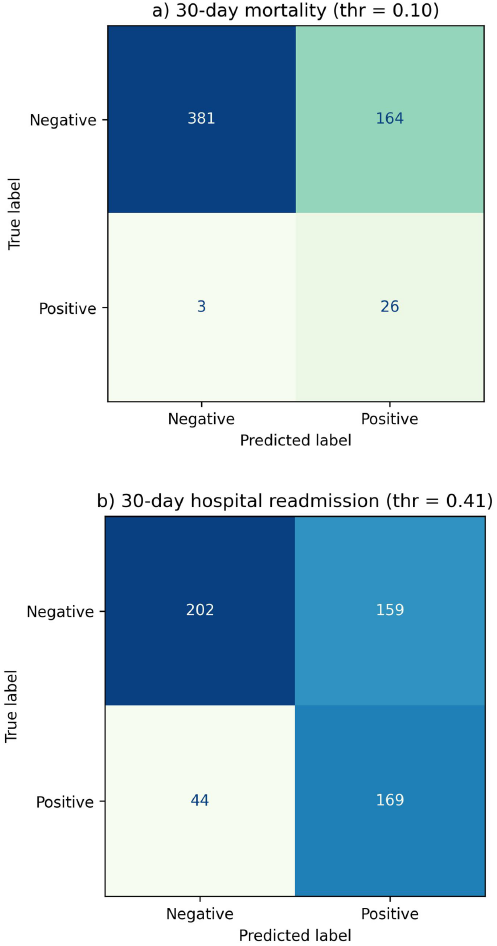
Confusion matrices for (a) 30-day mortality and (b) 30-day readmission LightGBM models on the test set, evaluated at a threshold of 0.10 and 0.41, respectively. Cell values are counts; axes show true vs. predicted labels.

**Figure 4.**
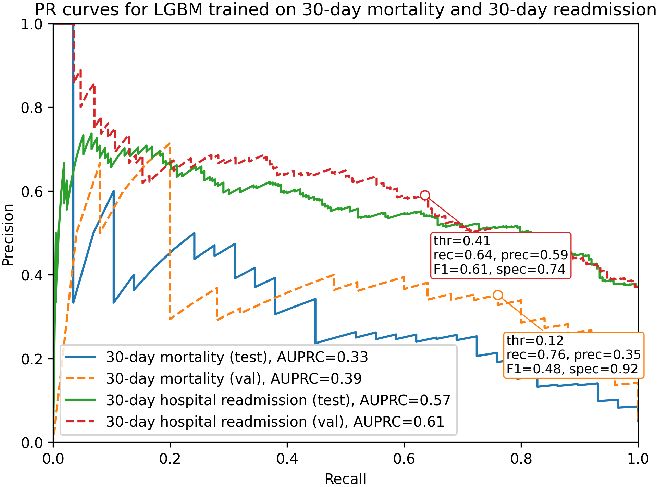
Precision–recall (PR) curves for the LightGBM model predicting 30-day mortality and 30-day hospital readmission. Solid lines show test-set performance (mortality AUPRC = 0.33; readmission AUPRC = 0.57), and dashed lines show validation-set performance (mortality AUPRC = 0.39; readmission AUPRC = 0.61). Callouts indicate validation-selected operating thresholds (thr) chosen to maximize the F1 score on the precision–recall curve (mortality thr = 0.12; readmission thr = 0.41), with corresponding recall, precision, F1 score, and specificity reported.

### 3.3. Feature importance for HF risk prediction

#### Validating previously identified features

Due to its performance on the test set, LightGBM was chosen for model explanation through SHAP analysis. Tree-based models such as GBMs have the advantage of providing feature importance data, which allows for identification of key clinical features and thresholds. Figure 5 shows the top 20 features and their impact on model output for LightGBM models trained on 30-day mortality and 30-day hospital readmission of HF patients. Some of the features identified have been previously identified to be correlated with mortality and/or hospital readmission. For instance, serum albumin has been previously identified as a predictor of cardiovascular mortality [45], and the LightGBM model also identified it as a key feature for the risk prediction.

**Figure 5.**
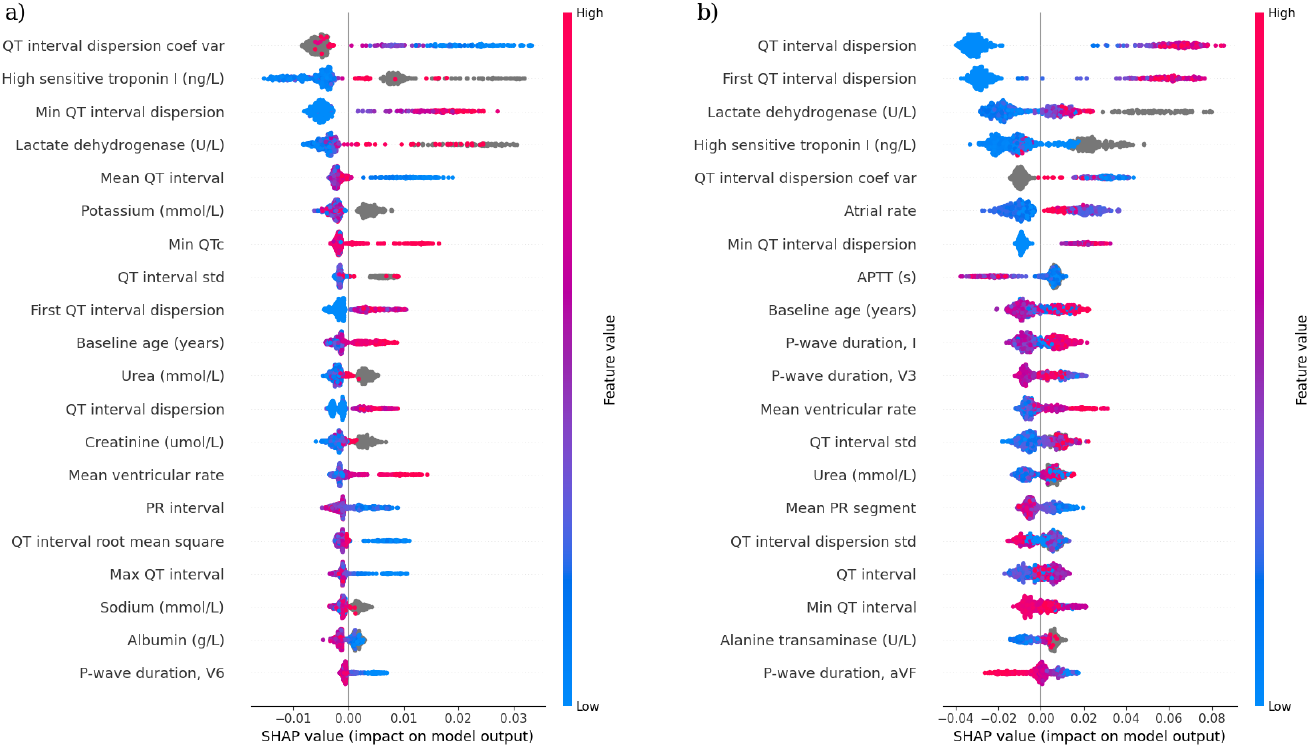
SHAP summary plot for the LightGBM models trained to predict (a) 30-day mortality and (b) 30-day hospital readmission. The plot shows the top 20 most impactful features on prediction and the distribution of each feature’s impact on the model output. Each point on the plot corresponds to an individual in the dataset. A negative SHAP value indicates reduced mortality risk, while a positive SHAP value indicates increased mortality risk.

The relationship between serum albumin levels and model prediction is demonstrated in Figure 6, and the model identified key thresholds of 35 g/L and 43 g/L identified for the 30-day mortality and hospital readmission separately; while previous study indicated that the normal reference range for albumin is 35-45 g/L, and a reduced serum albumin level is present in approximately 20% of all acute medical admissions [46]. Furthermore, LightGBM identifies QT interval as an important feature, which is also supported by previous literature [47]. As shown in Figure 6, QT intervals outside of the healthy range (350 to 450 ms for males and 360 to 460 ms for females) are associated with increased risk for mortality, particularly if the QT interval is below 350 ms.

**Figure 6.**
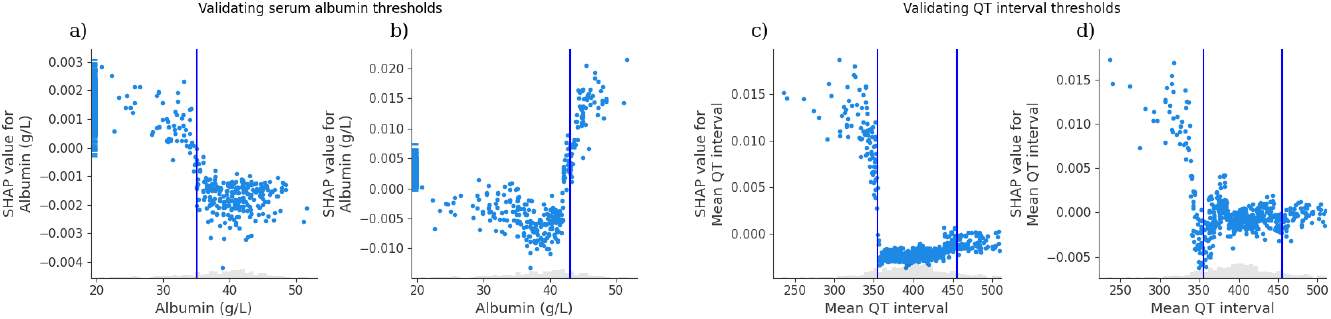
SHAP scatterplot showing the relationship between serum albumin levels and model prediction for (a) 30-day mortality and (b) 30-day hospital readmission. SHAP scatterplot showing the relationship between QT interval levels and model prediction for (c) 30-day mortality and (d) 30-day hospital readmission. The vertical lines represent key clinical thresholds. g/L identified in previous studies to be indicators of increased risk.

#### Identifying less explored features

Meanwhile, some of the top 20 features identified by LightGBM are less well-studied as risk factors for mortality and hospital readmission for HF patients. These features included lactate dehydrogenase level and QT dispersion. For example, Figure 7a shows a positive relationship between lactate dehydrogenase level and 30-day mortality, with a key threshold of around 600 U/L. LightGBM also identified a positive relationship between QT dispersion and 30-day readmission.

**Figure 7.**
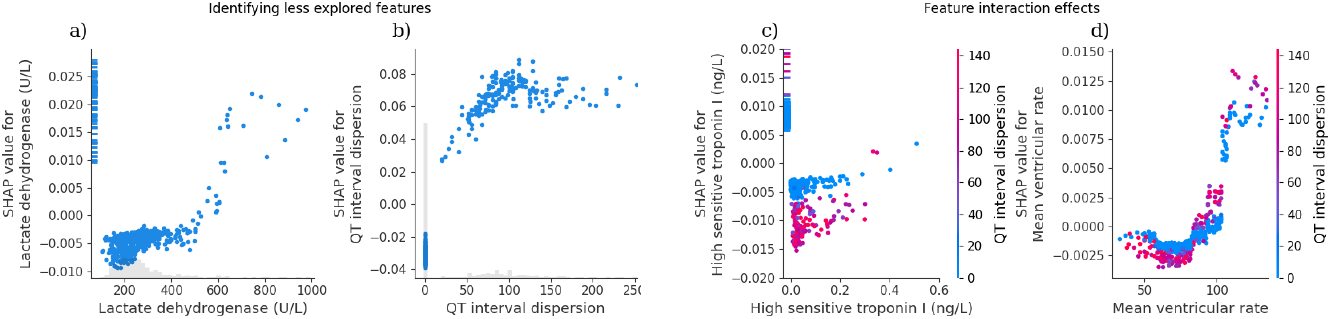
(a) SHAP scatterplot showing the relationship between lactate dehydrogenase levels and 30-day mortality U/L. (b) SHAP scatterplot showing the relationship between QT dispersion and 30-day hospital readmission. (c) Feature interaction effects between high sensitive troponin I levels and QT dispersion (d) Feature interaction effects between mean ventricular rate and QT interval dispersion.

#### Feature interaction effects

Furthermore, GBMs are effective at capturing feature interaction effects, giving insight into the complex relationship between the two features and the model outcome. For instance, Figure 7 demonstrate the interaction between the features high sensitive troponin level, mean ventricular rate, and QT dispersion. Figure 7c shows the interaction effect of high sensitive troponin levels with QT interval dispersion. Each point represents a patient, with color indicating QT interval dispersion values. The plot shows that for the same troponin I levels, patients with higher QT interval dispersion tend to have more negative SHAP values, indicating a stronger contribution to increased 30-day mortality risk. In contrast, patients with lower QT dispersion exhibit smaller or even SHAP effects at comparable troponin I concentrations. Figure 7d plots the interaction effect of mean ventricular rate and QT interval dispersion. In contrast to Figure 7c, this analysis shows that mean ventricular rate does not significantly interact with the effect of QT interval dispersion on 30-day mortality risk for patients.

The SHAP interaction values for every feature pair across the evaluation set are further visualized in Figure 8, where brighter cells indicate stronger two-way interactions in the model. The block structure highlights where the model most frequently exploits cross–feature effects: (*i*) dense and bright regions within the ECG modality (e.g., QT- and P–wave metrics with QRS morphology/dispersion features), (*ii*) within the laboratory test modality (e.g., electrolytes, lipids, troponin–related measures), and (*iii*) across ECG and laboratory test modalities. This suggests that while some ECG and lab features are partially redundant within their own modalities, combinations across ECG and laboratory domains are likely to interact and meaningfully impact predicted risk.

**Figure 8.**
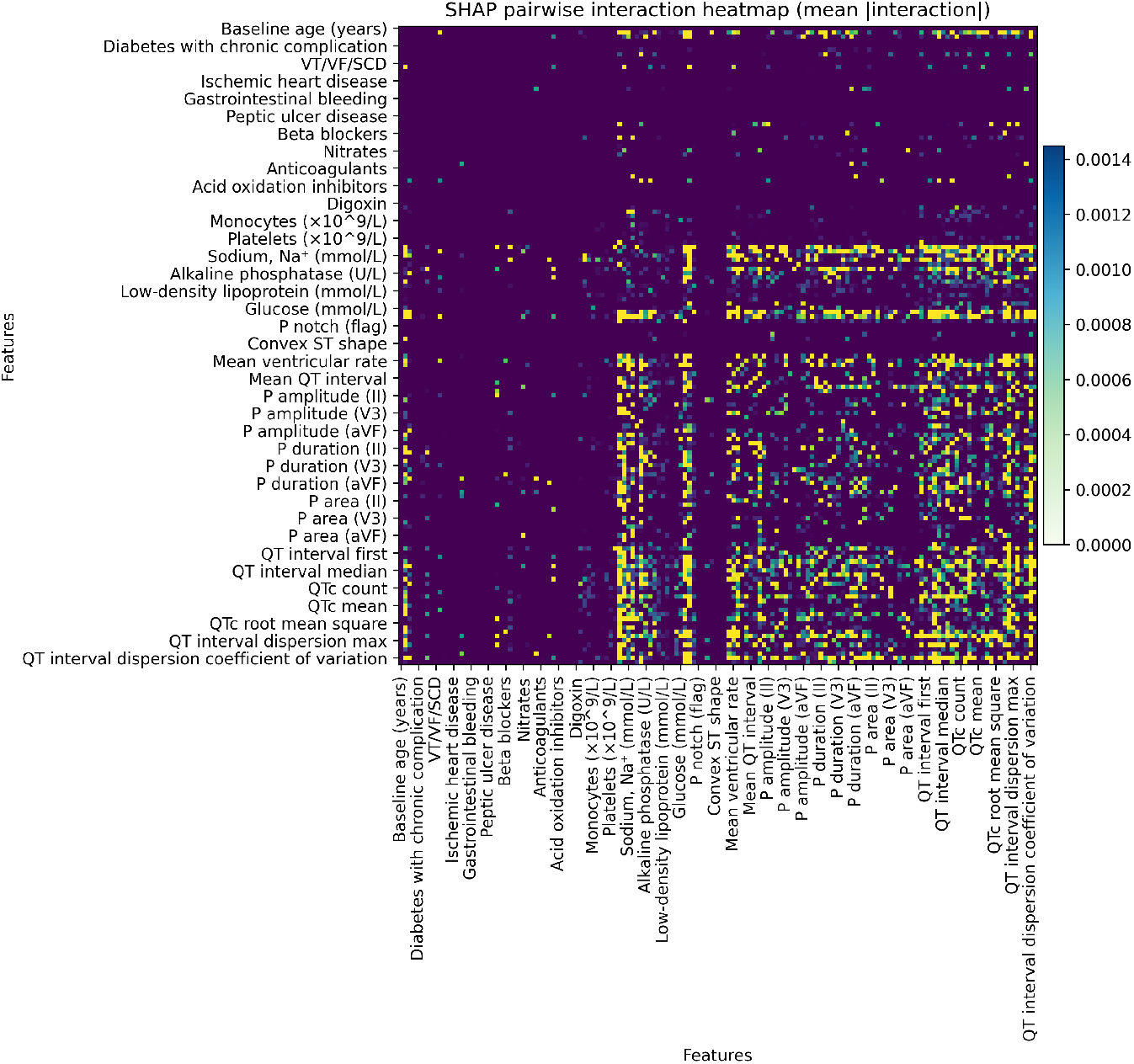
Supervised feature–feature distance heatmap. Each dot point in the image shows the pairwise supervised distance between model features (see Methods for metric definition). To preserve readability with 146 features, only every third feature label is shown on the axes. The color bar indicates the distance scale, and diagonal cells reflect self-comparisons.

### 3.4. Performance of multimodal data for HF risk prediction

One strength of our model is that it is trained on multimodal data, allowing for exploring interactions between different categories of patient data. The effect of multimodal data and interaction between these multimodal features on the model performance was investigated by grouping the features into different categories: (*i*) demographic characteristics and baseline comorbidities (denoted as Demo, or DB), (*ii*) medication prescriptions (Med, or MD), (*iii*) lab tests at admission (Lab, or LT), and (*iv*) ECG measurements (ECG). Table 3 shows results of HF risk prediction with combinations of the four feature categories, including the AUC, as well as specificity and sensitivity calculated at the operating point determined by Youden’s J statistic (i.e., the point maximizing sensitivity + specificity – 1) [43]. The first four rows show results for 30-day mortality and 30-day hospital readmission prediction for models trained on features from only one of the four feature categories. We report the predictive performance with different combinations of data modalities in the following subsections.

**Table 3.** Performance metrics for 30-day mortality and 30-day hospital readmission trained with different combinations of feature categories.

| Modalities | Feature Types | 30-day Mortality |  |  |  | 30-day Hospital Readmission |  |  |  |
| --- | --- | --- | --- | --- | --- | --- | --- | --- | --- |
|  |  | AUC | Sensitivity | Specificity | F1 | AUC | Sensitivity | Specificity | F1 |
| Single modality | Demo Features | 0.792 | 0.862 | 0.681 | 0.2530 | 0.646 | 0.465 | 0.798 | 0.5445 |
|  | Med Features | 0.547 | 0.793 | 0.349 | 0.0805 | 0.604 | 0.526 | 0.662 | 0.5489 |
|  | Lab Features | 0.802 | 1.000 | 0.508 | 0.2745 | 0.623 | 0.545 | 0.690 | 0.5469 |
|  | ECG Features | 0.809 | 0.793 | 0.717 | 0.1702 | 0.665 | 0.493 | 0.781 | 0.5390 |
| Two modalities | Demo + Med | 0.771 | 0.759 | 0.745 | 0.2162 | 0.685 | 0.610 | 0.709 | 0.5714 |
|  | Demo + Lab | 0.873 | 0.862 | 0.787 | 0.3279 | 0.666 | 0.667 | 0.598 | 0.5606 |
|  | Demo + ECG | 0.814 | 0.828 | 0.692 | 0.1778 | 0.671 | 0.671 | 0.629 | 0.5629 |
|  | Med + Lab | 0.806 | 1.000 | 0.484 | 0.2680 | 0.641 | 0.521 | 0.737 | 0.5523 |
|  | Med + ECG | 0.815 | 0.828 | 0.705 | 0.1778 | 0.682 | 0.765 | 0.571 | 0.6085 |
|  | Lab + ECG | 0.872 | 0.897 | 0.714 | 0.3500 | 0.698 | 0.765 | 0.548 | 0.5385 |
| Three modalities | Demo + Med + Lab | 0.858 | 1.000 | 0.631 | 0.3171 | 0.680 | 0.681 | 0.604 | 0.5771 |
|  | Demo + Med + ECG | 0.824 | 0.828 | 0.747 | 0.1702 | 0.689 | 0.723 | 0.596 | 0.5767 |
|  | Demo + Lab + ECG | 0.868 | 0.897 | 0.703 | 0.3636 | 0.699 | 0.723 | 0.598 | 0.5957 |
|  | Med + Lab + ECG | 0.863 | 0.897 | 0.703 | 0.3488 | 0.706 | 0.761 | 0.584 | 0.6076 |
| Four modalities | Demo + Med + Lab + ECG | **0.881** | 0.931 | 0.697 | 0.3768 | **0.709** | 0.793 | 0.562 | 0.5556 |

Note: Demo: demographic characteristics and baseline comorbidities (DB); Med: medication prescriptions (MD); Lab: Laboratory tests (LT); ECG: electrocardiogram-derived features. F1 scores are computed on the test set using thresholds selected on the validation set to maximize F1.

#### 3.4.1. Predictive performance with a single modality

As shown in Table 3, when using a single data modality for the HF risk prediction, ECG features achieved the highest performance (AUC: 0.809 for mortality, 0.665 for readmission), likely due to their ability to capture dynamic cardiac signals. Notably, Lab features also performed well on their own, likely due to their role as precise biomarkers of a patient’s physiological state (AUC: 0.802 for mortality, 0.623 for readmission). In contrast, Med features had the lowest performance with an AUC score of 0.547 for 30-day mortality prediction and 0.604 for 30-day readmission prediction. When considering F1 score, which reflects the balance between precision and recall under class imbalance, Lab features achieved the highest F1 among single-modality models for mortality (F1 = 0.27), while ECG features showed slightly lower F1 despite higher AUC, indicating that strong discrimination does not necessarily translate into optimal thresholded performance. For 30-day readmission, single-modality F1 scores were relatively similar across feature types (F1 0.54–0.55), suggesting limited gains from any single data source alone. Figures 9a and 10a demonstrate detailed comparison of model performance for predicting 30-day mortality and 30-day hospital readmission using features derived from a single data modality.

**Figure 9.**
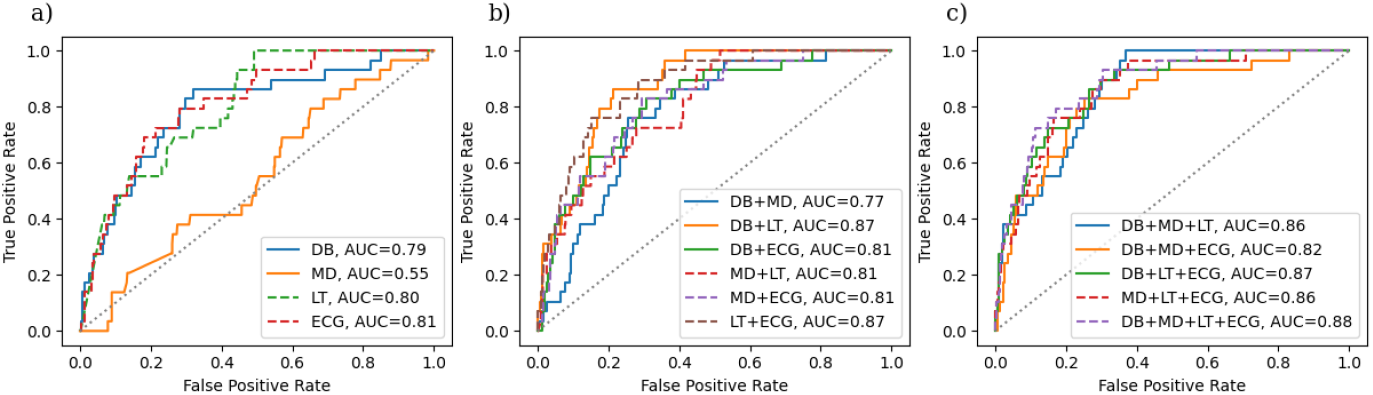
ROC curves for models trained for predicting 30-day mortality using (a) single feature modality, (b) combination of two feature modalities, and (c) combination of three or four feature modalities.

**Figure 10.**
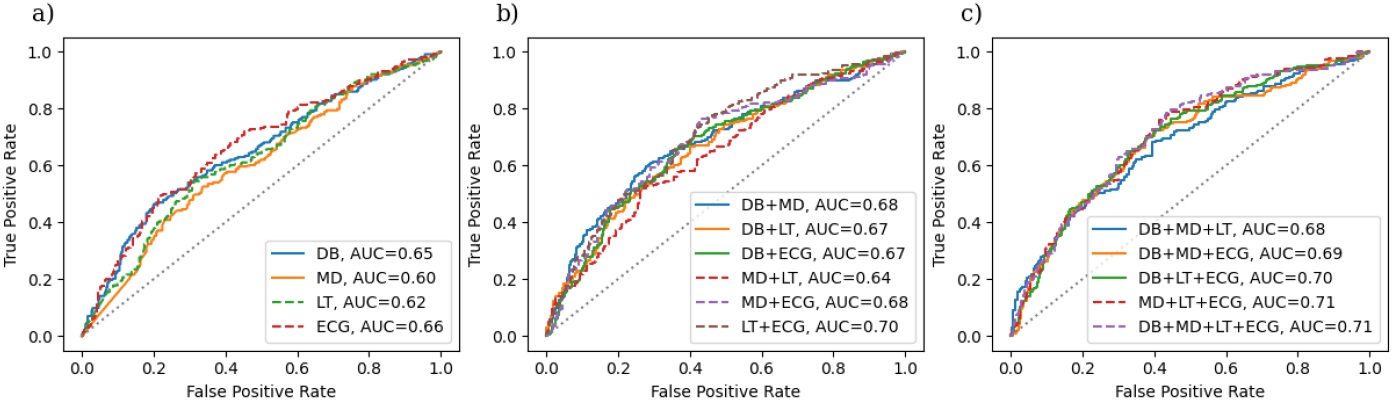
ROC curves for models trained for predicting 30-day hospital readmission using (a) single feature modality, (b) combination of two feature modalities, and (c) combination of three or four feature modalities.

**Figure 11.**
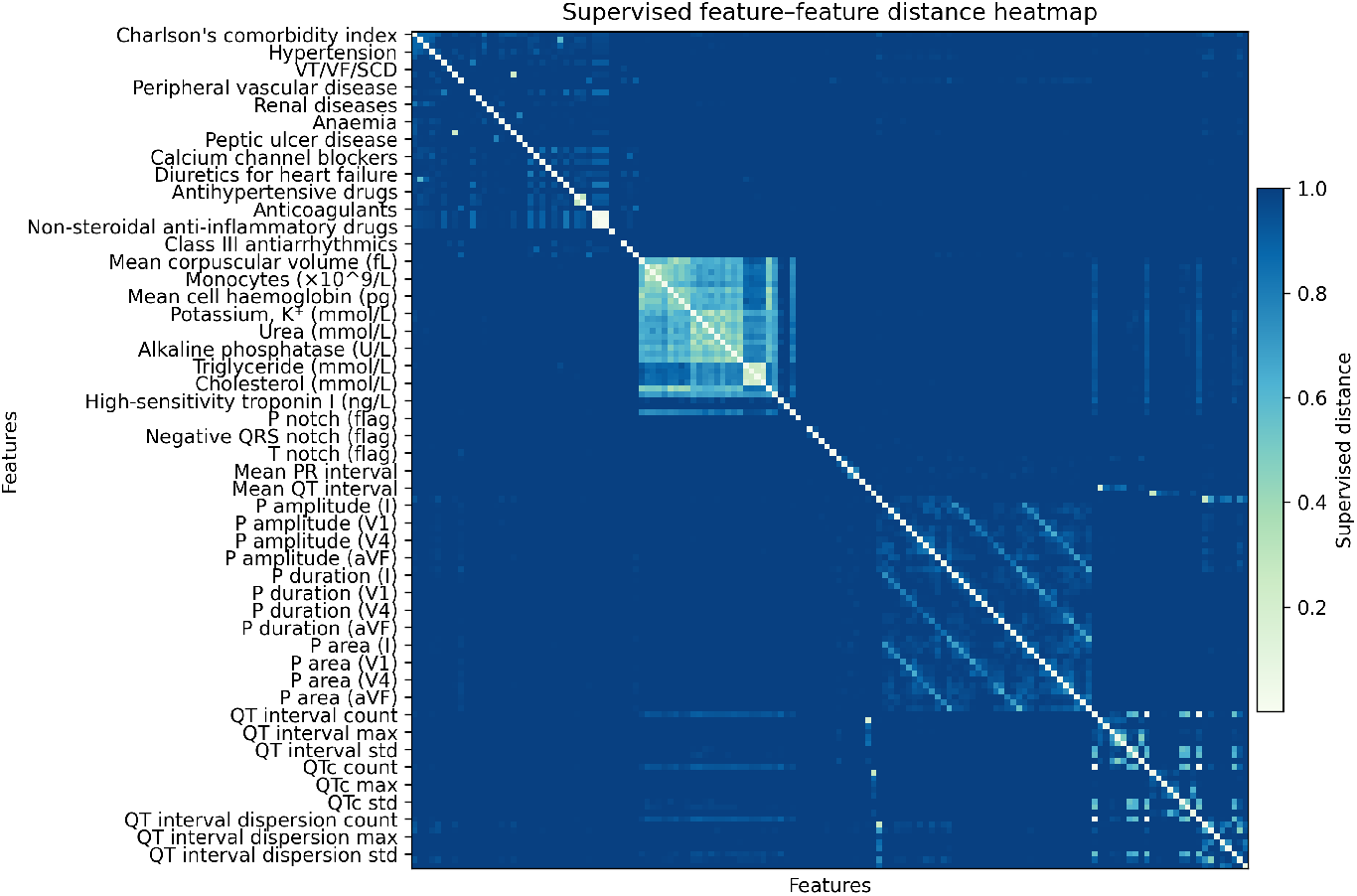
Supervised feature–feature distance heatmap. Each cell shows the pairwise supervised distance between model features (see Methods for metric definition). To preserve readability with 146 features, only every third feature label is shown on the axes. The color bar indicates the distance scale, and diagonal cells reflect self-comparisons.

#### 3.4.2. Predictive performance with two modalities

We explored six different combinations of two modalities of these features for the HF risk prediction. As shown in Table 3, when using two modalities of features, the combination of Lab and ECG features achieved high AUC scores of 0.872 and 0.698 for predicting 30-day mortality and 30-day hospital readmission respectively. This further validated our observation of the superior predictive performance when using Lab features or ECGs alone, as indicated in Section 3.4.1. Importantly, this performance gain was also reflected in F1 score, with Lab + ECG achieving the highest F1 among two-modality models for mortality (F1 = 0.35), indicating improved balance between sensitivity and precision at clinically relevant thresholds. Another interesting pattern is the comparison between Demo + Lab and Med + Lab. While both achieve high AUC values, Demo + Lab outperforms Med + Lab, particularly in specificity and sensitivity for predicting 30-day mortality. This difference is further supported by higher F1 scores for Demo + Lab (F1 = 0.33 vs. 0.27), highlighting the importance of demographic factors and baseline comorbidities in contextualizing biomarker information. The results also show that Lab + ECG performs slightly better than Demo + ECG. This reflects the strong prognostic value of laboratory tests that capture accurate patient health conditions, such as organ dysfunction, inflammation, and myocardial injury, which are directly linked to short-term outcomes in heart failure; while demographics provide important baseline risk (e.g., age, sex, comorbidities), offering a comprehensive profile for the risk prediction. The variations in performance underscore the importance of careful feature selection and the potential for over-fitting or noise when too many modalities are combined. Figures 9b and 10b provided detailed comparison of model performance for predicting 30-day mortality and 30-day hospital readmission using different combinations of two data modalities.

#### 3.4.3. Predictive performance with three modalities

We further investigated the predictive performance in HF risk using three modalities of these features. As shown in Table 3, integrating Lab features and ECGs either with Demo or Med features yields superior predictive performance compared to the other two feature configurations. These combinations also achieved consistently higher F1 scores for both outcomes, with Demo + Lab + ECG reaching an F1 of 0.36 for mortality and 0.60 for readmission, indicating improved robustness when additional contextual information is incorporated alongside physiological signals. We also observed that the model showed slightly decreased performance when comparing this combination Demo, Lab and ECG features to using only Lab and ECG, which may reflect added noise or redundancy in the data. This effect was more evident in AUC than F1, suggesting that while ranking performance may plateau, thresholded classification performance remains relatively stable. Figures 9c and 10c provided detailed comparison of model performance for predicting 30-day mortality and 30-day hospital readmission using different combinations of three modalities of features.

#### 3.4.4. Predictive performance with four modalities

As demonstrated in the above subsections, the comparison across different configurations of feature modalities indicates that incorporating more modalities generally improves the model’s performance in HF risk prediction. As shown in Table 3, using all four modalities achieved the highest AUC score, with 0.881 for 30-day mortality and 0.709 for 30-day hospital readmission respectively. This configuration also yielded the highest F1 score for mortality (F1 = 0.38) and competitive F1 performance for readmission (F1 = 0.56), suggesting that multimodal integration improves not only discrimination but also balanced classification performance when operating thresholds are selected on validation data. Figures 9c and 10c also included the detailed comparison of model performance for predicting 30-day mortality and 30-day hospital readmission using the four modalities of features.

### 3.5. Predictive performance of individual features

To analyze the predictive ability of individual features in each category, we trained reduced models using one feature from a single category and all features from the remaining three categories. Each reduced model includes a single feature from one category along with all features from the remaining categories. For instance, the reduced models for lab test category was trained using one lab test feature and all non-lab-related features. By doing so, these reduced models can shed insight on which features are the most influential in each category. Table 4 reports the feature rankings in both the original and reduced models, as well as the AUC scores indicating the predictive performance of each reduced model for 30-day mortality.

**Table 4.** Feature rankings and AUC performance for various features in original and reduced models.

| Feature | Feature ranking<br>(Original Model) | Feature ranking<br>(Reduced Model) | Predictive Performance of Reduced<br>Model (AUC) |
| --- | --- | --- | --- |
| **Baseline age (years)** | 10 | 5 | 0.8041 |
| Charlsons comorbidity index | 28 | 10 | 0.7789 |
| Anaemia | 131 | 23 | 0.7891 |
| Atrial fibrillation | 116 | 50 | 0.8003 |
| Diabetes with chronic complication | 131 | 58 | 0.8063 |
| **Nonsteroidal anti-inflammatory drugs** | 89 | 87 | 0.7963 |
| Antirheumatic drugs | 90 | 87 | 0.7963 |
| Antiplatelets | 131 | 88 | 0.8106 |
| ACEIARB | 77 | 90 | 0.7768 |
| Antihypertensive drugs | 110 | 99 | 0.8117 |
| **High sensitive troponin I (ng/L)** | 2 | 1 | 0.8142 |
| Lactate dehydrogenase (U/L) | 4 | 1 | 0.7929 |
| Potassium (mol/L) | 6 | 3 | 0.7522 |
| Urea (mmol/L) | 11 | 3 | 0.7503 |
| Creatinine (umol/L) | 13 | 3 | 0.7097 |
| **QT interval dispersion (coef var)** | 1 | 1 | 0.8171 |
| QT interval dispersion (min) | 3 | 1 | 0.8060 |
| QT interval (mean) | 5 | 2 | 0.7734 |
| QTc (mean) | 7 | 3 | 0.6941 |
| QT interval dispersion (first) | 9 | 3 | 0.7717 |

As shown in Table 4, the QT interval dispersion (coefficient of variation) is demonstrated as the most predictive feature in the ECG category, achieving the highest AUC score of 0.817. Therefore, the QT interval dispersion ranked the most important feature when representing its entire category. Similarly, other features such as QT interval dispersion (minimum) and QT interval (mean) demonstrate strong predictive performance. Similarly, high-sensitive troponin I and ACEIARB were the most predictive features in the lab tests and medication history category, respectively. In the demographics and baseline comorbidities category, age was the most predictive feature with a feature importance ranking of 5 in the reduced model compared to 10 in the original model. The fact that a single feature from each category retained significant predictive performance demonstrates some redundancy in features, and necessitating further exploring feature selection.

### 3.6. Exploring feature redundancy

The comparison of different feature modalities indicated that not all features contributed positively to the predictive performance, there are redundant or noisy features. To characterize dependencies among predictors, we computed a *supervised feature–feature distance* for exploring feature redundancy as follows [48].

For each ordered pair of features (*i, j*), we first trained a single–feature predictive model *f*_*i*_ using feature *x*_*i*_ to predict the clinical outcome *y*, yielding predictions *ŷ*_*i*_ = *f*_*i*_ (*x*_*i*_ ). We then trained a second single–feature model *g*_*j*←*i*_ using feature *x*_*j*_ to *mimic ŷ*_*i*_ by treating *ŷ*_*i*_ as the regression target. The supervised fit was quantified by

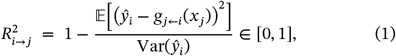

with values clipped below at 0.

This yields an *asymmetric* similarity: how well *x*_*j*_ can stand in for *x*_*i*_ . For visualization and clustering, we formed a conservative, *symmetric* distance

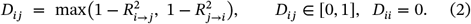

We refer to *D*_*ij*_ as the *supervised distance*: smaller values indicate higher redundancy (i.e., one feature can effectively substitute for the other), whereas larger values indicate complementary information.

The matrix **D** = [*D*_*ij*_] is demonstrated as a heatmap in Figure 11. Distinct light-colored blocks (low distance) appear within lab tests modality (e.g., electrolytes, urea, cholesterol, troponin) and within ECG measurements modality (e.g., interval and dispersion measures, P-wave/QRS morphology), indicating substantial within-group redundancy. These findings provide insights on redundancy-aware feature selection (e.g., clustering within blocks and retaining representatives, or aggregating correlated measures) to reduce noise and complexity while preserving predictive performance.

### 3.7. Subgroup analysis

To assess fairness and generalizability of the predictive model, we conducted subgroup analysis stratified by sex and age, evaluating performance for both 30-day mortality and 30-day hospital readmission (Figure 12). The performance score for each subgroup was estimated with 95% percentile bootstrap confidence intervals (1,000 resamples). The dashed vertical line in Figure 12 marks the overall AUC, while the dotted line at 0.5 corresponds to chance-level performance.

**Figure 12.**
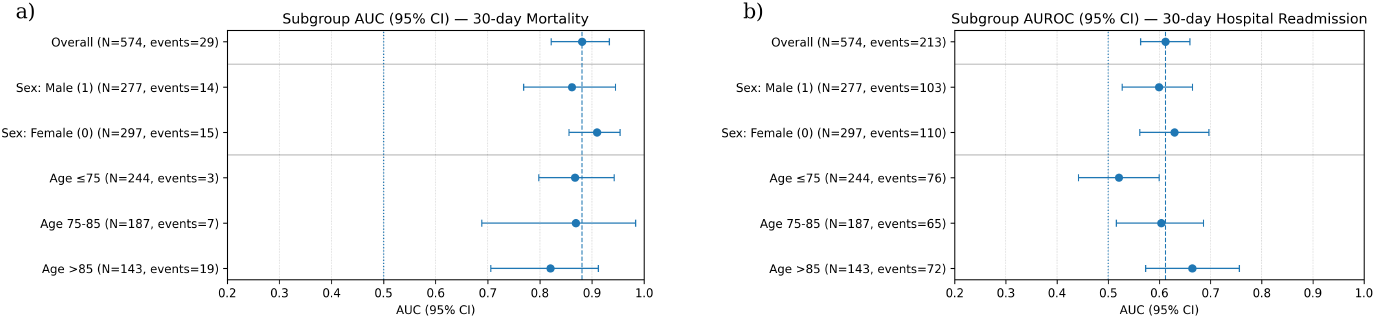
Subgroup discrimination on the test set for (a) 30-day mortality and (b) 30-day hospital readmission. Points show AUC with 95% percentile bootstrap CIs (1,000 resamples) for each subgroup; the dashed vertical line marks the overall AUC and the dotted line at 0.5 indicates chance performance. Horizontal separators delineate the overall, sex, and age sections. Labels include the numbers of patients and events.

As shown in Figure 12a, for 30-day mortality, stratified results indicated consistent model performance across subgroups, with similar AUCs for males (0.86) and females (0.88). Across age groups, the model maintained good discrimination in the 75-85 and >85 years age groups, although the ≤75 years group exhibited wide confidence intervals due to limited events (*n* =3), reflecting greater statistical uncertainty.

As shown in Figure 12b, for 30-day hospital readmission, the over-all AUC was lower than that for mortality prediction, consistent with the recognized challenges in accurately predicting readmission risk. Performance across sex subgroups was balanced, with overlapping confidence intervals between males and females for the readmission risk prediction. Age-based stratification showed comparable results, with modestly increased AUCs in older age groups. These sub-group analyses suggest that the models achieve stable performance across demographic groups. While we report subgroup discrimination, reliable subgroup-specific calibration and fairness metrics (e.g., PPV/NPV, TPR/FPR) could not be robustly estimated for certain strata—particularly younger age groups for 30-day mortality—due to limited event counts. Future validation using larger and more balanced cohorts will be essential to confirm equitable performance across different populations.

## 4. Discussion

This study presents a comprehensive and interpretable multimodal ML framework for predicting 30-day all-cause mortality and hospital readmission in HF patients using data from 2,868 individuals across 43 hospitals in Hong Kong. By integrating four distinct clinical data modalities, i.e., demographics, medications, laboratory tests, and ECG features, we systematically evaluated the predictive power of various ML model architectures and identified LightGBM as the top classifier with an AUC score of 0.881 for mortality and 0.709 for readmission. The use of SHAP analysis enabled robust model interpretation, revealing key predictors such as serum albumin, high-sensitivity troponin I, lactate dehydrogenase, and QT interval dispersion. These findings align with known pathophysiological mechanisms in mortality risk [45], [47], [49], while also highlight underrecognized biomarkers that may need further clinical investigation.

A major contribution of this work is the comprehensive ablation analysis across multimodal configurations, which quantified the incremental value of each data modality and their combinations. To the best of our knowledge, this is the first study to perform such a systematic evaluation of four clinical modalities in HF risk prediction. Our results demonstrate that laboratory tests and ECG features are the most informative individually and together achieve near optimal performance, suggesting that these two modalities may be adequate in resource constrained settings. Furthermore, the supervised feature distance analysis revealed substantial redundancy within laboratory tests and ECG features, indicating opportunities for model simplification through feature clustering or aggregation without sacrificing predictive accuracy.

Notably, combining all four modalities did not uniformly improve performance across metrics and modality configurations. Several factors may explain this pattern, including (i) feature redundancy and correlated predictors across modalities (e.g., overlapping clinical signal between laboratory tests, comorbidity indicators, and derived ECG features), (ii) noise introduced by higher-dimensional inputs relative to sample size, which can reduce generalization, and (iii) potential label noise or endpoint heterogeneity in real-world outcomes. Together, these results suggest that multimodal fusion is not guaranteed to be additive; in some settings, carefully selected subsets (e.g., Lab + ECG) may achieve near-optimal performance while offering lower complexity and improved deployability.

The strong performance of ECG features alone underscores the value of electrophysiological signals as dynamic indicators of cardiac instability. The identified ECG features also showed significant prognostic implications. For instance, QT dispersion has been showed to report worse clinical outcomes after arrest as it reflects the repolarization abnormalities [47]. It was suggested to be linked with cardiac and brain injury via sympathetic innervation of the left ventricle. Meanwhile, it was suggested that successful re-perfusion of ischemic cardiac tissues would decrease the QT dispersion [50]. The result demonstrated the model is capable of identifying pathophysiological factors that are contributing to adverse outcomes after HF. In addition, the interaction analysis showed meaningful cross modal effects particularly between lab and ECG domains, suggesting that combining biochemical and functional data enhances risk stratification. These interactions provide mechanistic hypotheses that could guide future research and support more personalized clinical decision making.

We acknowledge several limitations within this study. First, the population cohort used for this study was derived from a single and limited geographic region, which may affect generalizability of the developed model to other populations. Second, although we included multiple data modalities for the model development, medical images and longitudinal temporal patterns were not incorporated, these modalities may further improve the predictive performance of HF risks. Third, these ML models were trained on static clinical data, and did not account for in hospital changes or treatment responses, which may limit the performance of the developed framework in real-world clinical decision-making and care planning.

We additionally acknowledge that 30-day hospital readmission is influenced by social and health-system factors (e.g., discharge planning, outpatient follow-up access, caregiver support, socioeconomic context) that are not fully captured in structured EHR variables. Incorporating these factors, when available, may improve readmission prediction and interpretability in future work.

Our future research will explore several directions on advancing the health risk prediction and clinical translation. First, alternative model architectures will be investigated for improving the predictive performance. For instance, ensemble models may enhance predictive performance by combining multiple independently trained models; while transformer-based architectures such as TabTransformer [51] can be effective for tabular data, but remain underexplored in CVD outcomes prediction, leaving another route for future exploration. Second, generalizability of ML models is critical for real-world applications; we will explore external validation across diverse datasets from multiple institutions, enabling assessment of robustness across different populations and healthcare settings. Finally, prospective clinical trials in multicentre cohorts will be essential to validate these findings in healthcare setting, and evaluate the clinical utility and feasibility of model deployment.

## 5. Conclusion

This study presents a comprehensive multimodal machine learning framework for predicting 30-day all-cause mortality and hospital readmission in HF patients using four clinical data modalities: demographics, medications, laboratory tests, and ECGs. Our model achieved the predictive performance with an AUC of 0.881 for mortality and 0.709 for readmission. By performing a comprehensive analysis across four clinical modalities in HF risk modeling, our findings revealed that laboratory tests and ECG features contributed substantially to predictive performance, with their combination alone achieving near-optimal accuracy. We also identified clinically meaningful predictors such as serum albumin, lactate dehydrogenase, and QT interval dispersion, many of which reflect underlying pathophysiological mechanisms of cardiac dysfunction. These results underscore the importance of multimodal integration in enhancing risk prediction while offering transparency and practicality for real-world deployment. By balancing high performance, interpretability, and scalability, our framework supports personalized patient management and future implementation in diverse healthcare settings.

## Data Availability

All data produced in the present study are available upon reasonable request to the authors.

**Appendix Table 1.** Baseline and clinical characteristics of patients with heart failure stratified by Charlson’s standard comorbidity index. * for p≤ 0.05, ** for p ≤ 0.01, *** for p ≤ 0.001, IQR: interquartile range, ACEI: angiotensin-converting-enzyme inhibitors, ARB: angiotensin II receptor blockers. ^ indicates the variable distribution difference among four subgroups of patients by Charlson’s standard comorbidity index.

| Characteristics | All (N=2868)<br>Median (IQR);N or Count(%) | Charlson 0 (N=33)<br>Median (IQR);N or<br>Count(%) | Charlson 1-3 (N=608)<br>Median (IQR);N or<br>Count(%) | Charlson 4-6 (N=1528)<br>Median (IQR);N or Count(%) | Charlson 7+ (N=699)<br>Median (IQR);N or<br>Count(%) | P value^ |
| --- | --- | --- | --- | --- | --- | --- |
| **Outcomes** |  |  |  |  |  |  |
| 30-day mortality | 155(5.40%) | 0(0.00%) | 9(1.48%) | 73(4.77%) | 73(10.44%) | <0.0001*** |
| 30-day CVD mortality | 155(5.40%) | 0(0.00%) | 9(1.48%) | 73(4.77%) | 73(10.44%) | <0.0001*** |
| 30-day hospital readmission | 1065(37.13%) | 15(45.45%) | 201(33.05%) | 536(35.07%) | 313(44.77%) | 0.0016** |
| 30-day ASCVD hospital readmission | 408(14.22%) | 6(18.18%) | 70(11.51%) | 208(13.61%) | 124(17.73%) | 0.0104* |
| 30-day HF readmission | 207(7.21%) | 6(18.18%) | 43(7.07%) | 105(6.87%) | 53(7.58%) | 0.7532 |
| 30-day AF readmission | 169(5.89%) | 1(3.03%) | 17(2.79%) | 77(5.03%) | 74(10.58%) | <0.0001*** |
| 30-day stroke/TIA readmission | 47(1.63%) | 0(0.00%) | 1(0.16%) | 16(1.04%) | 30(4.29%) | <0.0001*** |
| 30-day AMI readmission | 93(3.24%) | 1(3.03%) | 18(2.96%) | 57(3.73%) | 17(2.43%) | 0.2196 |
| 30-day VT/VF/SCD readmission | 25(0.87%) | 0(0.00%) | 6(0.98%) | 9(0.58%) | 10(1.43%) | 0.1152 |
| 30-day IHD readmission | 121(4.21%) | 1(3.03%) | 15(2.46%) | 68(4.45%) | 37(5.29%) | 0.1486 |
| 30-day PVD readmission | 4(0.13%) | 0(0.00%) | 0(0.00%) | 1(0.06%) | 3(0.42%) | 0.0763 |
| **Demographics** |  |  |  |  |  |  |
| Sex (Male=1) | 1373(47.87%) | 18(54.54%) | 355(58.38%) | 687(44.96%) | 313(44.77%) | 0.2767 |
| Baseline age, years | 77.38(66.92-84.51);n=2868 | 42.67(33.59-46.64);n=33 | 60.63(53.74-68.02);n=608 | 79.18(72.69-85.4);n=1528 | 83.1(77.53-87.79);n=699 | <0.0001*** |
| Prior comorbidities |  |  |  |  |  |  |
| Charlson's comorbidity index | 5.0(4.0-6.0);n=2868 | 0.0(0.0-0.0);n=33 | 3.0(2.0-3.0);n=608 | 5.0(4.0-6.0);n=1528 | 8.0(7.0-9.0);n=699 | <0.0001*** |
| Diabetes without chronic complication | 848(29.56%) | 0(0.00%) | 69(11.34%) | 363(23.75%) | 416(59.51%) | <0.0001*** |
| Diabetes with chronic complication | 263(9.17%) | 0(0.00%) | 8(1.31%) | 54(3.53%) | 201(28.75%) | <0.0001*** |
| Hypertension | 1306(45.53%) | 3(9.09%) | 131(21.54%) | 670(43.84%) | 502(71.81%) | <0.0001*** |
| Heart failure | 2228(77.68%) | 14(42.42%) | 355(58.38%) | 1219(79.77%) | 640(91.55%) | 0.0005*** |
| Acute myocardial infarction | 281(9.79%) | 1(3.03%) | 30(4.93%) | 141(9.22%) | 109(15.59%) | <0.0001*** |
| VT/VF/SCD | 46(1.60%) | 0(0.00%) | 11(1.80%) | 27(1.76%) | 8(1.14%) | 0.3563 |
| Stroke/transient ischemic attack | 445(15.51%) | 0(0.00%) | 14(2.30%) | 165(10.79%) | 266(38.05%) | <0.0001*** |
| Atrial fibrillation | 825(28.76%) | 7(21.21%) | 155(25.49%) | 432(28.27%) | 231(33.04%) | 0.0385* |
| Peripheral vascular disease | 19(0.66%) | 0(0.00%) | 0(0.00%) | 3(0.19%) | 16(2.28%) | <0.0001*** |
| Ischemic heart disease | 1010(35.21%) | 2(6.06%) | 135(22.20%) | 514(33.63%) | 359(51.35%) | <0.0001*** |
| Hyperlipidaemia | 52(1.81%) | 0(0.00%) | 10(1.64%) | 17(1.11%) | 25(3.57%) | 0.0002*** |
| Renal diseases | 356(12.41%) | 0(0.00%) | 15(2.46%) | 84(5.49%) | 257(36.76%) | <0.0001*** |
| Liver diseases | 59(2.05%) | 0(0.00%) | 10(1.64%) | 20(1.30%) | 29(4.14%) | <0.0001*** |
| Gastrointestinal bleeding | 353(12.30%) | 0(0.00%) | 33(5.42%) | 179(11.71%) | 141(20.17%) | <0.0001*** |
| Anaemia | 433(15.09%) | 0(0.00%) | 30(4.93%) | 180(11.78%) | 223(31.90%) | <0.0001*** |
| Chronic obstructive pulmonary disease | 427(14.88%) | 1(3.03%) | 19(3.12%) | 214(14.00%) | 193(27.61%) | <0.0001*** |
| Cerebrovascular disease | 538(18.75%) | 0(0.00%) | 24(3.94%) | 210(13.74%) | 304(43.49%) | <0.0001*** |
| Peptic ulcer disease | 241(8.40%) | 0(0.00%) | 13(2.13%) | 94(6.15%) | 134(19.17%) | <0.0001*** |
| Cancer | 150(5.23%) | 0(0.00%) | 3(0.49%) | 33(2.15%) | 114(16.30%) | <0.0001*** |
| **Medications** |  |  |  |  |  |  |
| ACEI/ARB | 1517(52.89%) | 4(12.12%) | 259(42.59%) | 800(52.35%) | 454(64.94%) | <0.0001*** |
| Calcium channel blockers | 1300(45.32%) | 4(12.12%) | 180(29.60%) | 718(46.98%) | 398(56.93%) | <0.0001*** |
| Beta blockers | 1406(49.02%) | 8(24.24%) | 257(42.26%) | 759(49.67%) | 382(54.64%) | 0.0525 |
| Diuretics for hypertension | 335(11.68%) | 1(3.03%) | 61(10.03%) | 187(12.23%) | 86(12.30%) | 0.6484 |
| Diuretics for heart failure | 1173(40.89%) | 5(15.15%) | 184(30.26%) | 646(42.27%) | 338(48.35%) | 0.0038** |
| Anti-diabetic drugs | 1297(45.22%) | 5(15.15%) | 246(40.46%) | 616(40.31%) | 430(61.51%) | <0.0001*** |
| Nitrates | 801(27.92%) | 1(3.03%) | 78(12.82%) | 452(29.58%) | 270(38.62%) | <0.0001*** |
| Antihypertensive drugs | 535(18.65%) | 1(3.03%) | 57(9.37%) | 300(19.63%) | 177(25.32%) | <0.0001*** |
| Statins and fibrates | 866(30.19%) | 0(0.00%) | 153(25.16%) | 443(28.99%) | 270(38.62%) | <0.0001*** |
| Lipid-lowering drugs | 680(23.70%) | 0(0.00%) | 109(17.92%) | 351(22.97%) | 220(31.47%) | <0.0001*** |
| Anticoagulants | 556(19.38%) | 1(3.03%) | 136(22.36%) | 299(19.56%) | 120(17.16%) | 0.1748 |
| Antiplatelets | 1188(41.42%) | 0(0.00%) | 141(23.19%) | 637(41.68%) | 410(58.65%) | <0.0001*** |
| Anti-rheumatic drugs | 1172(40.86%) | 0(0.00%) | 139(22.86%) | 629(41.16%) | 404(57.79%) | <0.0001*** |
| Non-steroidal anti-inflammatory drugs | 1172(40.86%) | 0(0.00%) | 139(22.86%) | 629(41.16%) | 404(57.79%) | <0.0001*** |
| Acid oxidation inhibitors | 37(1.29%) | 0(0.00%) | 4(0.65%) | 17(1.11%) | 16(2.28%) | 0.0141* |
| Class I antiarrhythmics | 9(0.31%) | 0(0.00%) | 3(0.49%) | 5(0.32%) | 1(0.14%) | 0.5915 |
| Class III antiarrhythmics | 121(4.21%) | 0(0.00%) | 30(4.93%) | 61(3.99%) | 30(4.29%) | 1 |
| Class IV antiarrhythmics | 276(9.62%) | 0(0.00%) | 50(8.22%) | 153(10.01%) | 73(10.44%) | 0.4887 |
| Digoxin | 360(12.55%) | 2(6.06%) | 87(14.30%) | 189(12.36%) | 82(11.73%) | 0.5481 |
| **Complete blood counts** |  |  |  |  |  |  |
| Mean corpuscular volume, fL | 92.4(87.85-95.9);n=1394 | 90.4(88.3-93.3);n=17 | 91.4(86.95-95.4);n=336 | 92.6(88.3-96.0);n=737 | 92.5(87.35-96.4);n=304 | 0.9092 |
| Eosinophil, x10^9/L | 0.1(0.09-0.2);n=1207 | 0.1(0.1-0.2);n=15 | 0.1(0.1-0.2);n=272 | 0.1(0.07-0.2);n=640 | 0.1(0.08-0.22);n=280 | 0.7507 |
| Lymphocyte, x10^9/L | 1.2(0.8-1.7);n=1211 | 1.6(1.0-2.2);n=15 | 1.4(0.99-1.9);n=274 | 1.2(0.8-1.6);n=641 | 1.1(0.7-1.4);n=281 | <0.0001*** |
| Monocyte, x10^9/L | 0.5(0.4-0.7);n=1211 | 0.6(0.4-0.62);n=15 | 0.5(0.4-0.7);n=274 | 0.51(0.4-0.7);n=641 | 0.5(0.4-0.7);n=281 | 0.1901 |
| Neutrophil, x10^9/L | 5.0(3.8-7.0);n=1211 | 4.6(3.45-6.35);n=15 | 4.7(3.57-6.7);n=274 | 5.0(3.8-6.9);n=641 | 5.4(4.2-8.1);n=281 | 0.0003*** |
| White cell count, x10^9/L | 7.2(5.8-9.0);n=1394 | 7.0(5.8-7.67);n=17 | 7.1(5.7-8.6);n=336 | 7.2(5.8-8.8);n=737 | 7.4(6.1-9.8);n=304 | 0.0076** |
| Mean cell haemoglobin, pg | 31.4(29.6-32.9);n=1394 | 31.2(29.7-32.1);n=17 | 31.35(29.5-32.8);n=336 | 31.5(29.9-32.9);n=737 | 31.45(29.5-32.9);n=304 | 0.696 |
| Platelet, x10^9/L | 195.0(153.0-239.0);n=1393 | 211.0(189.0-274.0);n=17 | 202.0(165.0-244.5);n=336 | 192.0(151.0-238.0);n=736 | 192.0(148.0-238.5);n=304 | 0.1567 |
| Red blood count, x10^12/L | 3.9(3.38-4.4);n=1394 | 4.33(3.9-4.61);n=17 | 4.27(3.76-4.64);n=336 | 3.85(3.38-4.34);n=737 | 3.55(3.1-4.14);n=304 | <0.0001*** |
| **Liver and renal functions** |  |  |  |  |  |  |
| K/Potassium, mmol/L | 4.2(3.8-4.54);n=1666 | 4.1(4.0-4.5);n=21 | 4.2(3.9-4.5);n=454 | 4.2(3.8-4.6);n=867 | 4.2(3.8-4.6);n=324 | 0.8392 |
| Albumin, g/L | 39.65(35.9-42.7);n=1544 | 44.4(41.0-45.4);n=20 | 42.0(38.85-44.4);n=416 | 39.3(35.6-42.2);n=794 | 37.2(33.05-40.1);n=314 | <0.0001*** |
| Na/Sodium, mmol/L | 140.4(138.2-142.4);n=1667 | 139.6(138.0-141.3);n=21 | 140.7(138.8-142.0);n=454 | 140.5(138.3-142.5);n=867 | 139.8(137.1-142.7);n=325 | 0.0035** |
| Urea, mmol/L | 7.8(5.8-11.98);n=1667 | 5.75(4.8-7.1);n=21 | 6.7(5.2-9.15);n=454 | 7.8(5.98-11.53);n=867 | 11.0(7.5-19.7);n=325 | <0.0001*** |
| Protein, g/L | 72.5(68.0-76.4);n=1528 | 72.8(69.75-76.55);n=20 | 74.0(70.2-77.8);n=413 | 72.2(67.85-76.0);n=786 | 70.8(65.8-75.4);n=309 | <0.0001*** |
| Creatinine, umol/L | 108.0(84.0-152.0);n=1667 | 83.0(60.0-93.0);n=21 | 92.0(77.0-117.0);n=454 | 108.0(86.0-143.5);n=867 | 155.0(112.0-258.0);n=325 | <0.0001*** |
| Alkaline phosphatase, U/L | 77.0(62.0-99.0);n=1544 | 65.5(53.5-79.0);n=20 | 72.5(59.0-93.0);n=416 | 77.0(62.0-97.0);n=794 | 89.0(69.0-110.5);n=314 | <0.0001*** |
| Alanine transaminase, U/L | 19.0(14.0-28.0);n=1465 | 27.0(17.0-36.0);n=17 | 21.0(16.0-30.0);n=389 | 19.0(14.0-26.0);n=756 | 18.0(13.0-26.0);n=303 | 0.0006*** |
| Bilirubin, umol/L | 10.7(7.75-15.0);n=1539 | 11.0(8.5-14.5);n=20 | 11.0(8.0-16.0);n=416 | 10.95(7.75-15.25);n=792 | 10.0(7.0-13.1);n=311 | 0.0004*** |
| **Lipid profiles** |  |  |  |  |  |  |
| Triglyceride, mmol/L | 1.19(0.88-1.73);n=1049 | 1.18(0.9-1.31);n=11 | 1.25(0.94-1.9);n=315 | 1.16(0.85-1.69);n=546 | 1.16(0.88-1.66);n=177 | 0.2397 |
| Low-density lipoprotein, mmol/L | 2.29(1.8-2.9);n=1017 | 2.4(2.36-2.71);n=11 | 2.36(1.95-3.1);n=306 | 2.29(1.74-2.85);n=531 | 2.12(1.72-2.69);n=169 | 0.0139* |
| High-density lipoprotein, mmol/L | 1.25(1.0-1.54);n=1025 | 1.4(1.2-1.51);n=11 | 1.22(1.0-1.5);n=311 | 1.26(1.0-1.57);n=533 | 1.2(1.0-1.5);n=170 | 0.3989 |
| Total cholesterol, mmol/L | 4.3(3.6-5.0);n=1049 | 4.4(4.16-4.88);n=11 | 4.4(3.73-5.2);n=315 | 4.28(3.6-4.99);n=546 | 4.1(3.5-4.67);n=177 | 0.0041** |
| **Glucose tests** |  |  |  |  |  |  |
| HbA1c, g/dL | 11.8(10.4-13.4);n=1394 | 13.0(11.0-14.3);n=17 | 12.9(11.3-14.1);n=336 | 11.8(10.5-13.2);n=737 | 10.9(9.5-12.25);n=304 | <0.0001*** |
| Glucose, mmol/L | 6.3(5.4-8.22);n=1340 | 5.35(4.65-6.38);n=16 | 5.9(5.24-7.7);n=348 | 6.3(5.4-8.0);n=709 | 7.0(5.8-9.72);n=267 | <0.0001*** |
| **Other examinations** |  |  |  |  |  |  |
| High sensitive troponin-I, ng/L | 0.03(0.02-0.07);n=1935 | 0.12(0.02-0.59);n=15 | 0.02(0.01-0.05);n=402 | 0.03(0.02-0.06);n=1055 | 0.05(0.02-0.1);n=463 | <0.0001*** |
| Lactate dehydrogenase, U/L | 238.0(195.0-304.0);n=2453 | 338.5(248.0-448.5);n=26 | 248.0(200.0-325.5);n=503 | 234.0(193.0-298.0);n=1346 | 237.0(195.5-290.5);n=578 | 0.3409 |
| APTT, second | 33.5(29.9-39.9);n=1046 | 36.2(32.95-39.7);n=14 | 36.9(31.65-43.15);n=280 | 32.9(29.6-38.6);n=542 | 32.2(29.4-37.3);n=210 | 0.0007*** |
| **ECG measurements** |  |  |  |  |  |  |
| pnotchflag | 1066(37.16%) | 14(42.42%) | 221(36.34%) | 568(37.17%) | 263(37.62%) | 0.8789 |
| qrsdeltaflag | 17(0.59%) | 0(0.00%) | 6(0.98%) | 4(0.26%) | 7(1.00%) | 0.1855 |
| Positive qrsnotchflag | 608(21.19%) | 6(18.18%) | 107(17.59%) | 341(22.31%) | 154(22.03%) | 0.6556 |
| Negative qrsnotchflag | 425(14.81%) | 7(21.21%) | 91(14.96%) | 233(15.24%) | 94(13.44%) | 0.3402 |
| Convex stshape | 139(4.84%) | 1(3.03%) | 30(4.93%) | 72(4.71%) | 36(5.15%) | 0.758 |
| Concave stshape | 141(4.91%) | 0(0.00%) | 23(3.78%) | 67(4.38%) | 51(7.29%) | 0.0022** |
| tnotchflag | 802(27.96%) | 10(30.30%) | 177(29.11%) | 419(27.42%) | 196(28.04%) | 1 |
| atrialrate | 75.0(61.0-98.0);n=2867 | 92.0(70.0-112.0);n=33 | 77.0(64.0-99.0);n=608 | 74.0(60.0-97.0);n=1527 | 76.0(61.0-98.0);n=699 | 0.5922 |
| meanventrate | 77.0(67.0-90.0);n=2867 | 88.0(73.0-103.0);n=33 | 78.0(69.0-91.0);n=608 | 76.0(66.0-89.0);n=1527 | 77.0(66.0-90.0);n=699 | 0.7568 |
| meanprint | 176.0(157.0-200.0);n=2867 | 157.0(142.0-186.0);n=33 | 170.0(154.0-191.5);n=608 | 177.0(158.0-200.0);n=1527 | 180.0(161.0-205.0);n=699 | 0.0006*** |
| meanprseg | 79.0(64.0-99.0);n=2867 | 73.0(60.0-90.0);n=33 | 76.0(62.0-93.0);n=608 | 79.0(64.0-100.0);n=1527 | 82.0(66.0-104.0);n=699 | 0.0036** |
| meanqrsdur | 91.0(82.0-109.0);n=2867 | 93.0(78.0-111.0);n=33 | 91.0(84.0-107.0);n=608 | 92.0(82.0-110.0);n=1527 | 90.0(82.0-108.0);n=699 | 0.1199 |
| meanqtint | 393.0(360.0-428.0);n=2867 | 351.0(330.0-390.0);n=33 | 382.5(354.5-413.0);n=608 | 395.0(362.0-430.0);n=1527 | 400.0(364.0-440.0);n=699 | <0.0001*** |
| meanqtc | 444.0(420.0-472.0);n=2867 | 435.0(414.0-469.0);n=33 | 438.0(415.0-465.0);n=608 | 444.0(420.0-471.0);n=1527 | 452.0(426.0-476.5);n=699 | <0.0001*** |
| qtintdispersion | 0.0(0.0-72.0);n=2867 | 0.0(0.0-0.0);n=33 | 0.0(0.0-0.0);n=608 | 0.0(0.0-68.0);n=1527 | 60.0(0.0-112.0);n=699 | <0.0001*** |
| pamp_I | 0.07(0.01-0.1);n=2867 | 0.1(0.06-0.12);n=33 | 0.07(0.02-0.1);n=608 | 0.07(0.02-0.1);n=1527 | 0.06(0.01-0.09);n=699 | 0.002** |
| pamp_II | 0.09(0.03-0.13);n=2867 | 0.11(0.04-0.17);n=33 | 0.11(0.04-0.15);n=608 | 0.09(0.03-0.13);n=1527 | 0.08(0.01-0.12);n=699 | <0.0001*** |
| pamp_III | 0.06(0.0-0.1);n=2867 | 0.06(-0.04-0.1);n=33 | 0.07(0.0-0.11);n=608 | 0.06(0.0-0.1);n=1527 | 0.06(0.0-0.08);n=699 | 0.0008*** |
| pamp_V1 | 0.04(-0.05-0.07);n=2867 | 0.04(0.0-0.11);n=33 | 0.05(0.0-0.08);n=608 | 0.04(-0.06-0.07);n=1527 | 0.04(-0.06-0.07);n=699 | 0.0022** |
| pamp_V2 | 0.05(0.0-0.08);n=2867 | 0.04(0.0-0.09);n=33 | 0.05(0.01-0.09);n=608 | 0.05(0.0-0.08);n=1527 | 0.04(-0.04-0.07);n=699 | <0.0001*** |
| pamp_V3 | 0.06(0.01-0.08);n=2867 | 0.06(0.0-0.11);n=33 | 0.07(0.03-0.09);n=608 | 0.06(0.01-0.08);n=1527 | 0.05(0.0-0.08);n=699 | <0.0001*** |
| pamp_V4 | 0.06(0.01-0.08);n=2867 | 0.07(0.03-0.1);n=33 | 0.06(0.03-0.09);n=608 | 0.06(0.01-0.08);n=1527 | 0.05(0.01-0.07);n=699 | <0.0001*** |
| pamp_V5 | 0.05(0.01-0.08);n=2867 | 0.07(0.03-0.1);n=33 | 0.06(0.03-0.09);n=608 | 0.05(0.01-0.08);n=1527 | 0.05(0.01-0.07);n=699 | <0.0001*** |
| pamp_V6 | 0.05(0.01-0.08);n=2867 | 0.06(0.03-0.1);n=33 | 0.06(0.02-0.08);n=608 | 0.05(0.01-0.07);n=1527 | 0.04(0.01-0.07);n=699 | <0.0001*** |
| pamp_aVF | 0.07(0.02-0.11);n=2867 | 0.08(0.01-0.13);n=33 | 0.08(0.03-0.12);n=608 | 0.07(0.03-0.11);n=1527 | 0.06(0.01-0.09);n=699 | <0.0001*** |
| pamp_aVL | 0.01(-0.05-0.06);n=2867 | 0.04(-0.04-0.09);n=33 | 0.0(-0.05-0.06);n=608 | 0.01(-0.05-0.06);n=1527 | 0.01(-0.04-0.06);n=699 | 0.2132 |
| pamp_aVR | -0.07(-0.1-0.0);n=2867 | -0.09(-0.13-0.0);n=33 | -0.08(-0.11--0.02);n=608 | -0.07(-0.1--0.02);n=1527 | -0.06(-0.09-0.0);n=699 | <0.0001*** |
| pdur_I | 81.0(56.0-100.0);n=2867 | 78.0(62.0-98.0);n=33 | 84.0(64.0-98.0);n=608 | 81.0(56.0-100.0);n=1527 | 80.0(52.0-100.0);n=699 | 0.2453 |
| pdur_II | 92.0(68.0-110.0);n=2867 | 93.0(76.0-104.0);n=33 | 96.0(72.0-110.5);n=608 | 92.0(68.0-108.0);n=1527 | 92.0(65.0-112.0);n=699 | 0.5408 |
| pdur_III | 76.0(54.0-94.0);n=2867 | 80.0(68.0-96.0);n=33 | 79.0(60.0-96.0);n=608 | 74.0(52.0-92.0);n=1527 | 72.0(52.0-92.0);n=699 | 0.0558 |
| pdur_V1 | 52.0(39.0-76.0);n=2867 | 48.0(40.0-68.0);n=33 | 52.0(40.0-72.0);n=608 | 52.0(38.0-76.0);n=1527 | 54.0(38.0-80.0);n=699 | 0.4262 |
| pdur_V2 | 62.0(44.0-84.0);n=2867 | 64.0(40.0-73.0);n=33 | 64.0(46.5-84.0);n=608 | 62.0(44.0-84.0);n=1527 | 60.0(43.0-84.0);n=699 | 0.4024 |
| pdur_V3 | 76.0(56.0-96.0);n=2867 | 78.0(54.0-88.0);n=33 | 77.0(56.0-96.0);n=608 | 72.0(56.0-96.0);n=1527 | 76.0(52.0-98.0);n=699 | 0.4917 |
| pdur_V4 | 82.0(57.5-104.0);n=2867 | 80.0(56.0-92.0);n=33 | 86.0(60.0-102.5);n=608 | 84.0(58.0-104.0);n=1527 | 78.0(56.0-100.0);n=699 | 0.0169* |
| pdur_V5 | 87.0(60.0-104.0);n=2867 | 74.0(52.0-92.0);n=33 | 88.0(62.0-104.0);n=608 | 88.0(62.0-105.0);n=1527 | 84.0(58.5-104.0);n=699 | 0.5041 |
| pdur_V6 | 88.0(62.0-105.0);n=2867 | 76.0(56.0-92.0);n=33 | 87.0(62.0-104.0);n=608 | 88.0(62.0-106.0);n=1527 | 88.0(62.0-108.0);n=699 | 0.6082 |
| pdur_aVF | 84.0(64.0-102.0);n=2867 | 84.0(68.0-94.0);n=33 | 85.0(62.0-104.0);n=608 | 84.0(64.0-100.0);n=1527 | 84.0(63.0-102.5);n=699 | 0.9047 |
| pdur_aVL | 64.0(44.0-86.0);n=2867 | 64.0(42.0-90.0);n=33 | 64.0(44.0-84.0);n=608 | 64.0(44.0-86.5);n=1527 | 64.0(44.0-84.0);n=699 | 0.1521 |
| pdur_aVR | 92.0(64.0-108.0);n=2867 | 88.0(64.0-102.0);n=33 | 92.5(68.5-107.0);n=608 | 92.0(65.0-108.0);n=1527 | 90.0(64.0-108.0);n=699 | 0.499 |
| parea_I | 0.6(0.0-1.0);n=2867 | 0.9(0.4-1.4);n=33 | 0.7(0.1-1.2);n=608 | 0.6(0.1-1.0);n=1527 | 0.5(0.0-0.9);n=699 | 0.0028** |
| parea_II | 1.0(0.1-1.7);n=2867 | 1.2(0.3-2.1);n=33 | 1.3(0.2-2.05);n=608 | 1.0(0.1-1.7);n=1527 | 0.8(0.0-1.5);n=699 | <0.0001*** |
| parea_III | 0.5(0.0-0.9);n=2867 | 0.5(-0.2-1.0);n=33 | 0.6(0.0-1.15);n=608 | 0.5(0.0-0.9);n=1527 | 0.4(0.0-0.8);n=699 | 0.0011** |
| parea_V1 | 0.2(-0.4-0.5);n=2867 | 0.3(0.0-0.8);n=33 | 0.3(0.0-0.5);n=608 | 0.2(-0.4-0.45);n=1527 | 0.1(-0.5-0.4);n=699 | 0.003** |
| parea_V2 | 0.3(0.0-0.6);n=2867 | 0.3(0.0-0.7);n=33 | 0.4(0.0-0.7);n=608 | 0.3(0.0-0.6);n=1527 | 0.2(-0.3-0.6);n=699 | 0.0004*** |
| parea_V3 | 0.5(0.0-0.9);n=2867 | 0.7(0.0-1.2);n=33 | 0.6(0.2-1.0);n=608 | 0.5(0.0-0.8);n=1527 | 0.4(0.0-0.8);n=699 | <0.0001*** |
| parea_V4 | 0.6(0.0-1.0);n=2867 | 0.7(0.2-1.3);n=33 | 0.7(0.1-1.1);n=608 | 0.6(0.0-0.9);n=1527 | 0.5(0.0-0.9);n=699 | <0.0001*** |
| parea_V5 | 0.6(0.0-1.0);n=2867 | 0.7(0.2-1.1);n=33 | 0.7(0.1-1.2);n=608 | 0.6(0.0-1.0);n=1527 | 0.5(0.0-0.9);n=699 | <0.0001*** |
| parea_V6 | 0.6(0.0-1.0);n=2867 | 0.7(0.1-1.1);n=33 | 0.7(0.1-1.1);n=608 | 0.6(0.0-1.0);n=1527 | 0.5(0.0-0.9);n=699 | <0.0001*** |
| parea_aVF | 0.7(0.1-1.2);n=2867 | 0.6(0.0-1.7);n=33 | 0.9(0.1-1.5);n=608 | 0.7(0.1-1.2);n=1527 | 0.6(0.0-1.0);n=699 | <0.0001*** |
| parea_aVL | 0.0(-0.3-0.5);n=2867 | 0.2(-0.1-0.6);n=33 | 0.0(-0.3-0.5);n=608 | 0.0(-0.3-0.5);n=1527 | 0.0(-0.3-0.5);n=699 | 0.5661 |
| parea_aVR | -0.7(-1.3-0.0);n=2867 | -1.0(-1.4--0.3);n=33 | -0.9(-1.5-0.0);n=608 | -0.7(-1.2-0.0);n=1527 | -0.6(-1.1-0.0);n=699 | 0.0017** |
| meanqtint_count | 5.0(2.0-11.0);n=2868 | 5.0(2.0-9.0);n=33 | 7.0(3.0-15.0);n=608 | 5.0(2.0-11.0);n=1528 | 3.0(1.0-7.0);n=699 | <0.0001*** |
| meanqtint_first | 393.0(360.0-428.0);n=2868 | 351.0(331.0-390.0);n=33 | 382.5(354.0-413.0);n=608 | 396.0(360.5-430.0);n=1528 | 400.0(368.0-440.0);n=699 | <0.0001*** |
| meanqtint_min | 354.0(320.0-389.0);n=2868 | 335.0(318.0-376.0);n=33 | 346.0(312.5-376.0);n=608 | 352.0(320.0-389.0);n=1528 | 364.0(326.5-400.0);n=699 | <0.0001*** |
| meanqtint_max | 440.0(400.0-483.0);n=2868 | 421.0(372.0-444.0);n=33 | 444.0(402.0-483.0);n=608 | 440.0(400.0-486.0);n=1528 | 440.0(395.0-480.0);n=699 | 0.1074 |
| meanqtint_mean | 397.5(369.55-427.11);n=2868 | 374.33(348.0-403.33);n=33 | 394.54(370.15-419.08);n=608 | 398.58(370.06-428.0);n=1528 | 399.5(370.0-434.5);n=699 | 0.0778 |
| meanqtint_median | 399.5(369.0-428.75);n=2868 | 370.0(343.0-404.0);n=33 | 393.0(368.25-419.5);n=608 | 400.0(369.25-431.25);n=1528 | 400.0(370.75-436.0);n=699 | 0.053 |
| meanqtint_std | 35.57(23.9-50.0);n=2330 | 28.43(20.8-36.87);n=27 | 35.0(24.41-48.07);n=539 | 36.38(24.26-51.51);n=1246 | 34.72(23.18-50.67);n=518 | 0.8935 |
| meanqtint_coef_variation | 1.19(1.04-1.37);n=2720 | 1.16(1.05-1.33);n=32 | 1.25(1.1-1.4);n=573 | 1.2(1.04-1.37);n=1443 | 1.15(1.0-1.3);n=672 | <0.0001*** |
| meanqtint_root_mean_square | 400.0(371.77-429.17);n=2868 | 374.42(348.6-403.68);n=33 | 395.87(371.25-422.01);n=608 | 401.09(372.0-430.27);n=1528 | 401.18(374.01-435.99);n=699 | 0.094 |
| meanqtc_count | 5.0(2.0-11.0);n=2868 | 5.0(2.0-9.0);n=33 | 7.0(3.0-15.0);n=608 | 5.0(2.0-11.0);n=1528 | 3.0(1.0-7.0);n=699 | <0.0001*** |
| meanqtc_first | 444.0(420.0-472.0);n=2868 | 435.0(413.0-460.0);n=33 | 438.0(416.0-465.5);n=608 | 444.0(420.0-472.0);n=1528 | 451.0(428.0-478.0);n=699 | <0.0001*** |
| meanqtc_min | 418.0(394.0-443.0);n=2868 | 414.0(389.0-433.0);n=33 | 409.0(386.0-431.5);n=608 | 418.5(395.0-443.0);n=1528 | 429.0(403.0-456.0);n=699 | <0.0001*** |
| meanqtc_max | 485.0(454.0-518.0);n=2868 | 478.0(445.0-510.0);n=33 | 487.0(453.0-522.0);n=608 | 486.0(455.0-519.0);n=1528 | 480.0(452.0-514.5);n=699 | 0.097 |
| meanqtc_mean | 451.33(430.5-472.55);n=2868 | 443.27(427.25-453.5);n=33 | 447.0(427.25-468.92);n=608 | 451.52(431.18-471.58);n=1528 | 455.0(433.58-476.89);n=699 | 0.0005*** |
| meanqtc_median | 452.0(431.0-474.0);n=2868 | 440.0(428.0-449.0);n=33 | 446.5(427.0-470.5);n=608 | 452.0(432.0-473.0);n=1528 | 456.5(434.25-477.0);n=699 | <0.0001*** |
| meanqtc_std | 27.83(18.38-39.36);n=2330 | 27.58(19.58-32.65);n=27 | 27.34(18.79-39.6);n=539 | 28.55(18.99-39.51);n=1246 | 26.3(16.62-38.35);n=518 | 0.0488* |
| meanqtc_coef_variation | 1.13(1.03-1.24);n=2718 | 1.14(1.03-1.22);n=32 | 1.16(1.07-1.27);n=573 | 1.13(1.03-1.24);n=1441 | 1.1(1.0-1.2);n=672 | <0.0001*** |
| meanqtc_root_mean_square | 452.37(432.44-473.95);n=2868 | 443.9(428.11-454.12);n=33 | 447.83(428.31-470.94);n=608 | 452.69(433.06-473.44);n=1528 | 456.67(435.06-478.01);n=699 | 0.0009*** |
| qtintdispersion_count | 5.0(2.0-11.0);n=2868 | 5.0(2.0-9.0);n=33 | 7.0(3.0-15.0);n=608 | 5.0(2.0-11.0);n=1528 | 3.0(1.0-7.0);n=699 | <0.0001*** |
| qtintdispersion_first | 0.0(0.0-72.0);n=2868 | 0.0(0.0-0.0);n=33 | 0.0(0.0-0.0);n=608 | 0.0(0.0-68.0);n=1528 | 56.0(0.0-112.0);n=699 | <0.0001*** |
| qtintdispersion_min | 0.0(0.0-34.5);n=2868 | 0.0(0.0-0.0);n=33 | 0.0(0.0-0.0);n=608 | 0.0(0.0-20.0);n=1528 | 0.0(0.0-72.5);n=699 | <0.0001*** |
| qtintdispersion_max | 116.0(0.0-192.0);n=2868 | 0.0(0.0-140.0);n=33 | 115.5(0.0-182.0);n=608 | 114.0(0.0-193.0);n=1528 | 120.0(0.0-193.5);n=699 | 0.0641 |
| qtintdispersion_mean | 57.76(0.0-96.0);n=2868 | 0.0(0.0-58.47);n=33 | 35.73(0.0-71.63);n=608 | 59.92(0.0-96.45);n=1528 | 81.33(0.0-113.52);n=699 | <0.0001*** |
| qtintdispersion_median | 52.0(0.0-93.0);n=2868 | 0.0(0.0-26.0);n=33 | 0.0(0.0-72.75);n=608 | 52.0(0.0-92.25);n=1528 | 80.0(0.0-108.0);n=699 | <0.0001*** |
| qtintdispersion_std | 46.09(8.49-67.28);n=2330 | 34.3(0.0-58.2);n=27 | 47.59(12.0-65.09);n=539 | 46.36(0.35-68.3);n=1246 | 42.43(14.37-65.37);n=518 | 0.8618 |
| qtintdispersion_coef_variation | 1.7(1.0-2.89);n=769 | 2.31(2.31-2.31);n=2 | 2.09(1.04-3.24);n=50 | 1.75(1.0-3.06);n=391 | 1.6(1.0-2.68);n=326 | 0.113 |
| qtintdispersion_root_mean_square | 75.3(0.0-111.73);n=2868 | 0.0(0.0-69.24);n=33 | 59.89(0.0-93.33);n=608 | 75.46(0.0-112.94);n=1528 | 92.0(0.0-124.14);n=699 | <0.0001*** |

## Notes

### Competing Interest Statement

The authors have declared no competing interest.

### Funding Statement

Acknowledged.

### Author Declarations

The project obtained ethics approval from the Chinese University of Hong Kong-New Territories East Cluster Clinical Research Ethics Committee for studies related to the use of electrocardiographies for risk stratification (Approval numbers: 2019.338, 2019.361, and 2019.422).

